# Explainable, personalised prediction of emergency readmission and mortality following hospitalisation in patients with heart failure

**DOI:** 10.64898/2026.07.15.26358201

**Authors:** B Gallego, L Huberts, J Yu, V Blake, L Jorm, L Liu, S-Y Ooi

## Abstract

**Background:** Unplanned emergency readmissions remain common following hospitalisation for heart failure (HF). Residual congestion, atrial fibrillation, frailty, and other comorbidities contribute to adverse outcomes after discharge. Identifying patients at high risk of readmission or death may help target post-discharge management.

**Methods:** We conducted a retrospective cohort study of patients hospitalised with HF in selected New South Wales hospitals who were discharged alive and not documented as receiving end-of-life care. Clinical, laboratory, medication, and text-derived variables extracted from electronic health records were used to develop predictive models and corresponding risk scores for emergency readmission and all-cause mortality within 180 days of discharge. Feature importance methods were used to identify key predictors and explain individual risk estimates. To illustrate model predictions while preserving patient privacy, we generated representative synthetic patient profiles by summarising the characteristics of groups of patients with similar predicted risk patterns and visualised the major contributors to their predicted risks using Shapley values.

**Results:** The study included 5,202 hospitalisations among 3,933 patients. Within 180 days of discharge, 45.2% of patients experienced at least one emergency readmission and 12.4% died. The most common causes of emergency readmission were recurrent HF, followed by atrial fibrillation, chest pain, and pneumonia. Predictive performance was moderate for emergency readmission (AUC 0.70; calibration slope 1.30) and good for mortality (AUC 0.84; calibration slope 1.01). Emergency readmission risk was primarily associated with greater prior healthcare utilisation, a higher number of active medical problems, high risk of falls, older age, and impaired kidney function. Mortality risk was most strongly associated with abnormal red blood cell distribution width, elevated blood urea, older age, and lower systolic blood pressure. A lower number of discharge medications, particularly cardiovascular therapies, was associated with a higher risk of emergency readmission and a lower risk of mortality. Representative synthetic patient profiles demonstrated heterogeneity in the factors contributing to predicted risks, illustrating the value of patient-level risk visualisation.

**Conclusions:** Predictive models identified clinically meaningful predictors of emergency readmission and mortality following HF hospitalisation. Patient-level visualisation of individual risk drivers may support more personalised post-discharge management.

## 1. Introduction

Heart failure (HF) is a complex clinical syndrome affecting approximately 1-2% of the adult population. Its prevalence is expected to surge 46% by 2030 due to population aging, increased prevalence of risk factors and improved survival.^1^

The post-discharge period following hospitalisation for heart failure remains a critical phase characterised by persistently high rates of adverse events. In the Patient-Centered Care Transitions in Heart Failure (PACT-HF) trial,^2,3^ nearly 50% of patients experienced a hospital readmission, emergency department presentation, or death within the first three months after discharge; by three years of follow-up, over 90% of patients had experienced at least one of these events. Analysis of the European Society of Cardiology Heart Failure registry indicates that residual congestion is present in approximately 31% of patients at discharge and is associated with worse subsequent outcomes.^4^

Observational evidence also suggests that outcomes after heart failure hospitalisation remain poor. A systematic review and meta-analysis of Australian data^5^ reported pooled all-cause readmission rates of 20% at 30 days and 56% at one year, with corresponding mortality rates of 8% at 30 days and 25% at one year. Similarly, a recent report by the Australian Institute of Health and Welfare^6^ reported 30-days emergency readmission rate of 23%, and 1-year emergency readmission and mortality rates of 64% and 24% respectively.

Multiple interventions have been proposed to address the high burden of adverse outcomes following hospitalisation for heart failure, with mixed evidence of effectiveness. The PACT-HF trial^2,3^ evaluated a comprehensive transitional care model incorporating nurse-led home visits, heart function clinic follow-up, self-care education, structured discharge summaries, and early physician review. While this intervention improved patient-reported outcomes, including discharge preparedness, quality of care transitions, and quality of life in the early post-discharge period, extended follow-up demonstrated no sustained reduction in all-cause mortality or hospitalisation over three years. In contrast, recent evidence from the Safety, Tolerability, and Efficacy of Rapid Optimization, Helped by NT-proBNP Testing, of Heart Failure Therapies (STRONG-HF) trial^7^ focused on early initiation and rapid up-titration of guideline-directed medical therapy before discharge, combined with frequent and closely monitored follow-up during the first six weeks after hospitalisation, resulting in significant reductions in readmission and mortality within six months from discharge.

Telemedicine and remote monitoring^8,9^ have been extensively evaluated as strategies to reduce adverse outcomes following heart failure hospitalisation, with heterogeneous results. Systematic reviews and meta-analyses suggest that non-invasive telemonitoring may be associated with reductions in all-cause mortality and heart failure hospitalisations in selected settings, particularly when monitoring data are linked to timely therapeutic action and medication optimisation. This is consistent with findings from the STRONG-HF trial. These findings suggest that remote monitoring is most likely to be effective when embedded within structured, treatment-focused care pathways, rather than as a standalone intervention. However, these interventions are often resource-intensive, and their scalability and cost-effectiveness remain uncertain.

Identification of patients at higher risk of adverse events can enable more efficient and targeted allocation of post-discharge resources. Improving risk stratification at the time of discharge therefore represents a critical opportunity to reduce potentially avoidable readmissions, optimise care transitions, and facilitate timely goals-of-care and end-of-life discussions where appropriate.

A substantial body of literature has examined prediction of 30-day readmission and/or mortality following hospital discharge for heart failure. A recent systematic review of machine-learning (ML) approaches for predicting all-cause emergency (unplanned) 30-day readmission^10^ identified marked heterogeneity across studies, with reported readmission rates ranging from 1.2% to 39.4%, wide variation in modelling approaches and discriminative performance, and infrequent reporting of calibration metrics. One of the largest and most methodologically rigorous studies in this area, using linked data from the ‘Get With The Guidelines–Heart Failure registry and Medicare claims’^11^, compared multiple ML and traditional statistical models for predicting 30-day all-cause readmission. Across models, discrimination was consistently modest, with C-statistics ranging from 0.57 to 0.62. Calibration analyses demonstrated reasonable agreement between predicted and observed risk in the mid-range of predicted probabilities, but miscalibration at the extremes. A smaller number of studies have examined predictive models beyond the 30-day window. A systematic review evaluating models including 90 and 180-day horizons^12^ found modest discrimination and limited calibration reporting across these extended endpoints.

This study focusses on explainable, patient-level risk stratification for predicting unplanned hospital readmission and all-cause mortality within 180 days of discharge among patients hospitalised with heart failure. Using the same follow-up horizon as STRONG-HF, we examine patients who have not been identified as approaching end of life and develop visual explanations of each patient’s predicted risk, highlighting the individual clinical factors contributing to that risk. The aim is to move beyond risk prediction alone towards interpretable, actionable information that can support personalised post-discharge care planning. We leverage a comprehensive CardiacAI electronic medical record dataset linked to population health data^13^, enabling incorporation of symptom information extracted from unstructured clinical notes, and inclusion of discharge medications, recognising their potential to modify post-discharge risk.

## 2. Methods

### 2.1. Data source

This cohort study utilised data from the Cardiac Analytics and Innovation (CardiacAI) Data Repository (version 1.2). CardiacAI^13^ prospectively collects and curates Electronic Medical Record (EMR) data from cardiovascular patients admitted to hospitals in two Local Health Districts in NSW. This data is then linked to the NSW Admitted Patient Data Collection (APDC), which contains records of all hospital admissions in NSW, and to the NSW Registry of Births, Deaths and Marriages (RBDM). The linkage is conducted by the Centre for Health Record Linkage (CHeReL).^14^ The EMR data includes patient demographics, ward movements, emergency triage, diagnoses, surgeries, medications, blood transfusions, vital signs, pathology, patient family and social history, as well as clinical notes.

### 2.2. Study population

The study cohort included patients >18 years of age admitted to seven hospitals in New South Wales (NSW), Australia between January-1st-2017, and March-31st-2021, with a diagnosis of Heart Failure (ICD-10-AM code I50), as both primary and secondary diagnosis. We chose to analyse the broader group of hospitalised people living with HF rather than limiting the study to patients hospitalised with a principal diagnosis of HF, since HF patients can be admitted for various reasons related to their disease.

Hospital encounters ending in patient death or patient being transferred to another hospital or to a different service were excluded from the cohort of index admissions but their Length of Stay (LOS) was taken into account as part of the subsequent admission. Also excluded were encounters with LOS<24 hours, often related to brief admissions for dialysis. Patients transferred to a hospice or already receiving palliative (end-of-life) care were also excluded, as they are already identified as requiring special consideration. These patients were identified using Palliative care diagnosis codes (ICD10: Z51.5, SNOMED:103735009, 305824005, 305686008, 441874000, 305496007, 1084981000168105, 719239007, 713673000), having a Palliative Care consult note, as well as using a rule-based concept extraction method in the clinical notes (see Supplementary material Table 1). End-of-Life care concept extraction has been previously validated against 600 annotated notes. There were 65 true positive notes and 535 true negative notes. The algorithm correctly identified 44 of the 65 positive notes and 528 of the 535 negative notes, corresponding to a precision of 86.3% and recall (sensitivity) of 67.7%.

### 2.3. Outcomes

The primary outcomes were at least one all-cause emergency readmission, and all-cause death within 180 days from the index discharge, identified through the linked administrative data (APDC and RBDM). A readmission was classified as emergency if it was recorded with a status of unplanned/urgent admission and involved admission within an emergency department with level three or higher urgency, not just minor cases.

### 2.4. Patient and admission characteristics

The following patient characteristics were analysed in the development dataset for inclusion in our predictive models.

### Patient demographics

Included age, sex, indigenous status, language spoken at home, country of birth, married status, financial class and postcode of residence. Financial class, which refers to the patient status in relation to healthcare payments (e.g. public patient or overseas private patient) had >50% missing values and was therefore excluded from the analysis. Postcode of residence was excluded due to small cell sizes. No other variables had missing data, though some included categories labelled “Other/Unknown”.

### Patient social and family history

We looked at information such as: patient lives alone, concern about social isolation, patient exercises, alcohol use, smoking status and patient is employed; all potentially relevant for the prediction of unplanned readmission. However, values for all these variables were largely missing (likely because this information comes from a form mostly used in outpatient settings). Only 15 reported items were present in at least 1% of admissions and considered in the analysis.

### Index admission characteristics

Included hospital, specialty, ward, admission type (mostly related to emergency vs. planned-non-emergency), and LOS. Only ward at admission had missing values, which were encoded as such.

### Hospital stays in the previous year

We calculated the cumulative length of stay (cumLOS) for all hospital admissions within the State during the year preceding the index admission. Because some index admissions did not have a full year of prior history available and the sample size was already small, we conducted a sensitivity analysis comparing three approaches: (i) using cumLOS as observed, (ii) adjusting cumLOS by the proportion of available history, and (iii) setting cumLOS to missing when available history was less than one year. Results were consistent across approaches and the cumLOS rate was selected as the potential predictor.

### Pathology

We included findings from the most prevalent pathology tests for cardiac patients, totalling 191 distinct test types within our heart failure cohort. For non-numerical variables, we took the last reported value during the admission. For numerical variables, we estimated the mean over all available measurements weighted by measurement time, with the latest measurement having the highest weight. We subsequently excluded test types from the analysis due to a missingness rate exceeding 90%, keeping 85 pathology measurements. Missingness was treated as informative and left un-imputed, enabling the tree-based model to handle missing values intrinsically.

### Vital signs

The most commonly performed vital signs measurements for cardiac patients were included, totalling 111 distinct types. For non-numerical variables, we took the last reported value during the admission. For numerical variables, we estimated the mean over all available measurements weighted by measurement time, with the latest measurement having the highest weight. We subsequently excluded vital sign measurements from the analysis due to a missingness rate exceeding 90%, keeping 39 measurement types. Missingness was treated as informative and left un-imputed, enabling the tree-based model to handle missing values intrinsically.

### Medications

We considered medication orders that started before discharge (including orders from previous admissions) and did not stop before discharge. Medications were classified using the Cerner Multum DNUM drug identifier, which groups medications at the ingredient (drug concept) level and resulted in 526 types, which were then grouped into 134 primary groups (see Supplementary material, Table 2). Each drug identifier was assigned 1 (at least one such medication at discharge) or 0. Medications groups not associated with cardiovascular disease prescribed to less than 1% of admissions were excluded from the analysis, leaving 54 medication types.

### Problems

Over 1,000 confirmed and active problems were reported of which only 16 appeared in at least 1% of admissions and were considered in the analysis.

### Surgeries

We looked at the primary procedure associated with the last surgery performed in each index admission and found a total of 155 distinct procedures. Most of them were performed in less than 1% of encounters and were subsequently excluded from the analysis, only ‘Echocardiogram transoesophageal and cardioversion’ remained. This is likely due to the fact that procedures taking place in private cardiac catheterisation laboratories were not available in our data set.

### Symptoms

Selected concepts related to symptoms from the Palliative care Outcome Scale (POS) (such as ‘dyspnoea’, ‘pain’, ‘depression’, ‘reduced activities of daily living’), as well as ‘frailty’ and ‘deterioration’, were extracted from all clinical notes. The number of notes in which a concept was found during each encounter was used as a proxy for the severity of the symptom, and therefore as a predictor. Also, concepts related to ‘preserved ejection fraction’ and ‘reduced ejection fraction’ were extracted and combined with ‘Heart failure with reduced ejection fraction’ reported under Problems, to build a HFrEF flag. Concept extraction took place using a rule-based method (see Supplementary material Table 1).

### 2.5. Data pre-processing

Data were randomly partitioned into a model development (training) set (80%) and a test set (20%), ensuring that each patient appeared in only one set to avoid data leakage. Since the sample size is relatively small, the partition was stratified by outcome. The test set was never used during model development.

Using the development dataset, categorical variables were one-hot encoded, continuous variables were standardised and all binary variables were mapped to 0/1. Predictor variables (with the exception of concepts extracted from clinical notes) were then filtered based on variance and univariate association with the outcome. Variables with low variability were excluded (variance < 0.01 for binary variables and < 0.001 for continuous variables). Binary, continuous, and categorical predictors were further excluded if their effect size fell below predefined thresholds (binary: risk difference<0.02, continuous: standardised mean difference<0.1, categorical: maximum category-specific risk difference<0.02) and their univariate out-of-sample AUC was < 0.55. For variables to be removed, the lack of univariate association had to hold for both emergency readmission and death.

### 2.6. Predictive algorithm and performance evaluation

Predictive models were developed using gradient-boosted decision trees implemented in the python library XGBoost^15^. Hyperparameters were optimised using the Optuna framework^16^, which applies Bayesian optimisation to efficiently explore the hyperparameter space. Optuna was applied with cross-validation that was stratified by patient and outcome to prevent information leakage across folds and to preserve outcome prevalence during tuning. To improve robustness and reduce variance, the final predictive model was defined as an ensemble obtained by averaging predictions across multiple cross-validated bootstrap resamples of the training data and changing random model initialisation seeds. Each model in the ensemble was trained using the optimal hyperparameter configuration identified by Optuna. Because bootstrapping was performed at the patient level rather than the encounter level, the resulting prediction intervals more accurately reflected between-patient uncertainty and avoided artificially narrow uncertainty estimates arising from correlated encounters within the same individual. Probability calibration was applied using isotonic regression and although it had a small improvement in calibration, it did not result in significant improvements (see Supplementary material Figure 1).

The predictive performance of the ensemble model was evaluated using a bootstrapping procedure on both the train and the test set, given the relatively small size of the test cohort. This allowed us to account for two sources of uncertainty: model uncertainty related to variations from the model and training data, and sampling uncertainty related to variations in the test population. Reported metrics included the Receiver Operating Characteristic (ROC) curve, the Area Under the Curve (AUC), the Brier Score, and the calibration curve and slope.

### 2.7. Feature importance and explanation of individual risk estimates

To quantify the contribution of individual features to model predictions, we used SHAP (SHapley Additive exPlanations) values using TreeSHAP.^17^ For each observation in the test set, feature importance was defined as the mean SHAP value computed across the bootstrapped training models. Because SHAP values are additive on the logit scale, averaging preserves the additive decomposition of the ensemble logit prediction. Global feature importance was estimated by averaging the absolute ensemble SHAP values across all observations in the test set.

To explain the predicted risk for an individual patient, cumulative SHAP contributions were first computed on the logit scale and then sequentially transformed to the probability scale, allowing feature effects to be displayed as incremental changes in predicted risk in a waterfall plot. The plot was anchored to the reported ensemble predicted probability (defined as the mean predicted probability across bootstrap models). Owing to the nonlinear logit-to-probability transformation, a residual arises between the transformed cumulative logit and the ensemble predicted probability, however, this residual was negligible (∼0.1%).

### 2.8. Alignment with TRIPOD conventions

This study follows the TRIPOD AI reporting guidelines. The checklist can be found in the Supplementary material.

## 3. Results

There were 7,292 hospital encounters among 5,367 patients with a heart failure (HF) diagnosis during the study period. After excluding encounters that ended in in-hospital death, transfer to another hospital, hospice, or service, as well as encounters involving patients previously identified as end-of-life and those with a length of stay <24 hours, 5,202 index admissions among 3,933 patients remained for analysis (Table 1).

**Table 1:** Cohort derivation flowchart.

| FLOWCHART | excl. | encs. | pats. |
| --- | --- | --- | --- |
| Initial cohort |  | 7292 | 5367 |
| Died in hospital | 388 |  |  |
|  |  | 6904 | 5080 |
| Transferred to a hospice | 9 |  |  |
|  |  | 6895 | 5074 |
| Transferred to other hospital/service | 1186 |  |  |
|  |  | 5709 | 4225 |
| Identified as EOL | 227 |  |  |
|  |  | 5482 | 4133 |
| LOS<24 hours | 280 |  |  |
| Final cohort |  | 5202 | 3933 |

| OUTCOMES | encs. (%) | pats. (%) |
| --- | --- | --- |
| **Within 30 days** |  |  |
| all cause death | 107(2.1%) | 104(2.6%) |
| all cause read. | 1458(28.0) | 1197(30.4) |
| all cause e. read. | 1149(22.1) | 940(23.9%) |
| **Within 180 days** |  |  |
| all cause death | 591(11.4%) | 488(12.4%) |
| all cause read. | 3169(60.9) | 2364(60.1) |
| all cause e. read. | 2362(45.4) | 1778(45.2) |

Within this cohort, 60.1% of patients experienced at least one hospital readmission and 45.2% experienced at least one emergency readmission within 180 days post-discharge. Figure 1 presents the most common principal diagnoses associated with these emergency readmissions. Although not being identified as end-of-life at baseline, 12.4% of patients died within 180 days of discharge.

**Figure 1:**
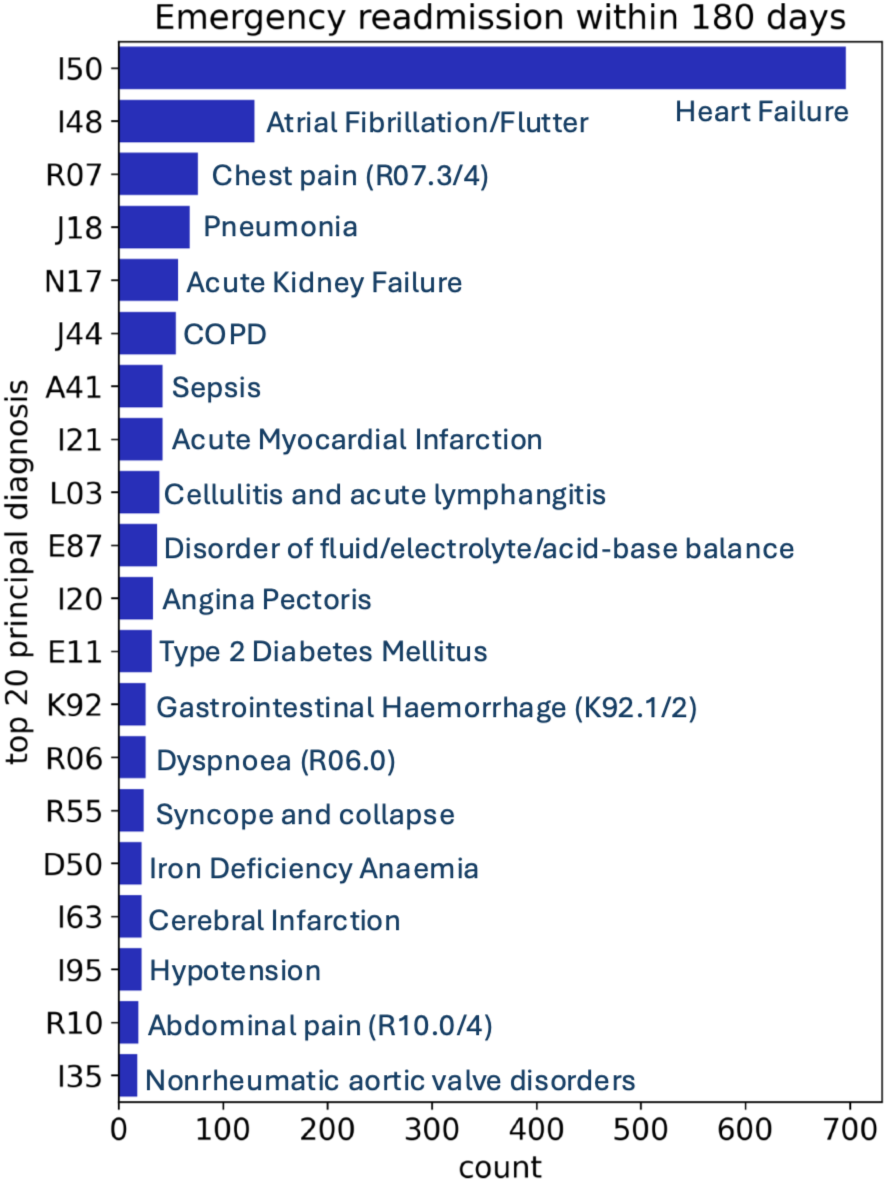
Principal diagnoses distribution of emergency readmissions within 180 days from discharge

Table 2 summarises the distribution of baseline patient characteristics for patients who experienced emergency readmission or death vs. those who did not. Given the large number of available variables, only selected pathology measures and vital signs are presented, focusing on those identified as most relevant for the predictive models. Likewise, only cardiovascular-related medication classes are reported. Overall, most demographic characteristics, laboratory measurements, and vital signs differed significantly between patients with and without emergency readmission or death. Age, cumulative length of stay, index admission length of stay, red cell distribution width, urea level and troponin had higher values in the emergency readmission and death groups; whereas lower renal function, albumin, lymphocytes and diastolic blood pressure were found in these groups. Being married or having a defacto partner was less common amongst patients who experienced readmission.

**Table 2:**
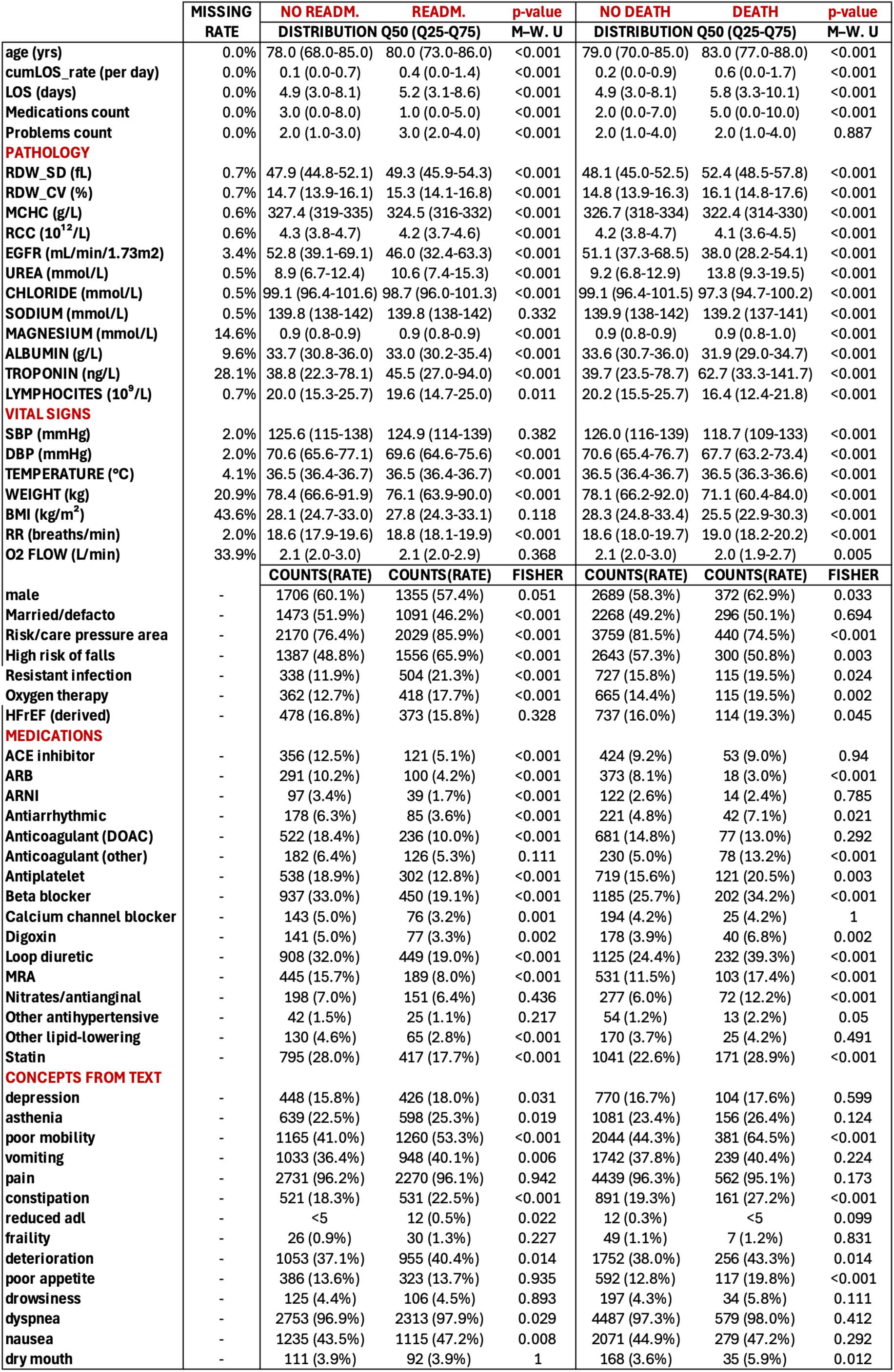
Distribution of selected baseline characteristics across four groups: No emergency readmission vs. emergency readmission and No death vs. death.

Patients who experienced an emergency readmission had higher number of problems and were generally prescribed fewer medications (including cardiovascular medications) at discharge; while patients who died had a larger number of prescriptions. Patients with emergency readmission had significantly more frequent mentions of poor mobility, vomiting, nausea and constipation. Among patients who died, poor mobility, constipation, and poor appetite were more commonly documented.

### 3.1. Models’ predictive performance

Despite the modest sample size, the predictive models demonstrated moderate performance for emergency readmission and good performance for death. For emergency readmission (Figure 2 – top panel) the area under the receiver operating characteristic curve (AUC) was 0.70 [0.66,0.73], the Brier score was 0.22 [0.21,0.23] and the calibration slope=1.30. Using the probability threshold associated with maximum geometric mean of sensitivity and specificity, 550 encounters (52%) in the test set were classified as at risk of emergency readmission and may represent candidates for intensified post-discharge monitoring. Amongst those, 321 (PPV=58%) were true positives and accounted for 68% of all emergency readmissions. This coverage can be increased by reducing the probability threshold at the cost of more false positives. For example, a threshold probability of 0.30, identifies 93% of all patients who experienced an emergency readmission, with the PPV reduced to 51%.

**Figure 2:**
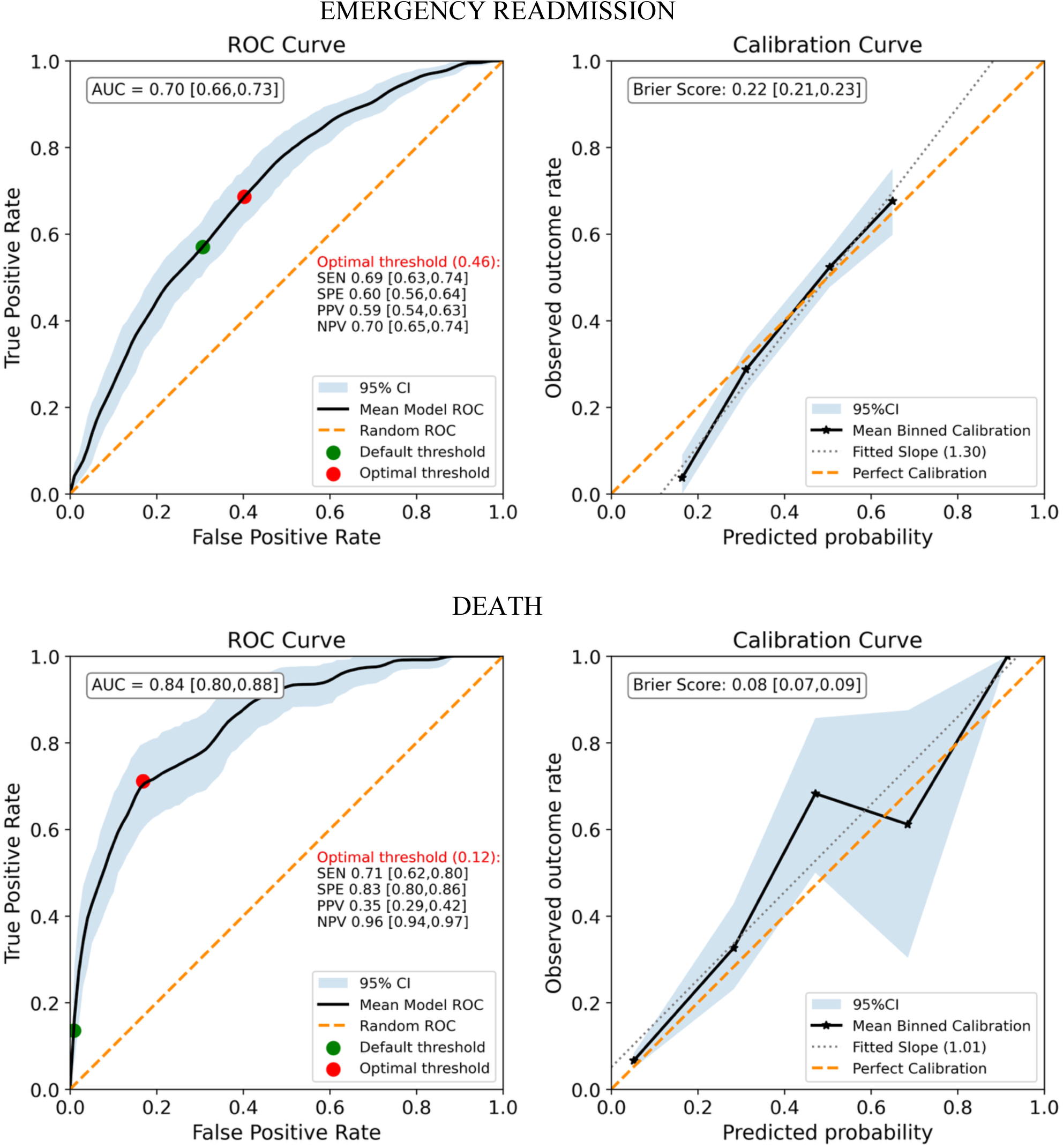
Receiver Operating Curves and calibration curves corresponding to the emergency readmission risk prediction model (top panel) and the mortality risk prediction model

For mortality (Figure 2 – lower panel), the AUC was 0.84 [0.80,0.88] with a Brier score of 0.08[0.07-0.09] and calibration slope of 1.01. Uncertainty around mortality risk estimates was substantial, likely reflecting the relatively low event rate, limited sample size, and the fact that these estimates represent residual risk after exclusion of patients recognised as end-of-life. At the threshold of maximum geometric mean of sensitivity and specificity, 239 encounters in the test set (23%) were classified as being at risk of dying within the next 180 days. Of those, only 84 were true positives (PPV=35%) but they represented 71% of all deaths.

### 3.2. Variables associated with predicted risk

Figure 3 presents SHAP summary plots illustrating the contribution of individual features to the models’ predictions. Features were ranked according to their mean absolute SHAP values across all observations in the test set. Each point represents a single observation; its horizontal position indicates the SHAP value (i.e., the feature’s impact on the prediction), while its colour represents the feature value from low (blue) to high (red). Features with wider SHAP value distributions exert a greater influence on model predictions. In general, red points with positive SHAP values (right) indicate that higher feature values increase the predicted risk, whereas red points with negative SHAP values (left) indicate that higher feature values decrease the predicted risk.

**Figure 3:**
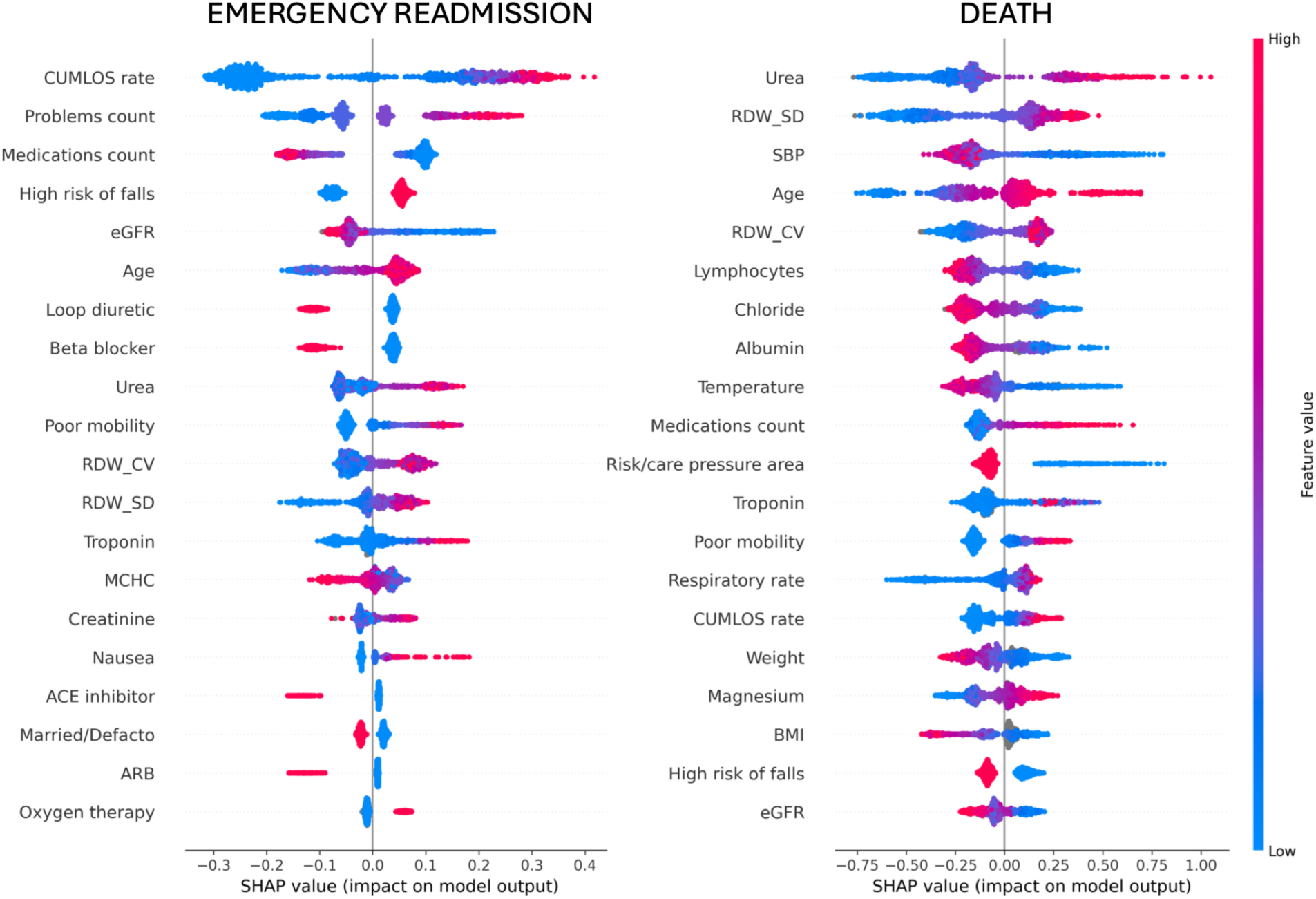
Top 20 predictors by feature importance as estimated using Shap values for emergency readmission (left) and death (right). Each point represents a single observation; its horizontal position indicates the SHAP value (i.e., the feature’s impact on the prediction), while its colour represents the feature value from low (blue) to high (red).

Emergency readmission risk was primarily associated with greater prior healthcare utilisation and a higher number of recorded active problems. A lower number of medications at discharge, particularly cardiovascular medications, was generally associated with an increased risk of emergency readmission but a lower risk of mortality. This pattern may partly reflect the competing risk of early death occurring before a hospital readmission. Figure 4 further examines the mean number and distribution of medications and documented active problems across patients grouped by their outcomes. Among patients who survived beyond 180 days, those who were not readmitted had higher medication counts than those who were readmitted. Patients who died within 180 days generally had higher medication counts than survivors, particularly among those who were not readmitted. Logistic regression analysis restricted to patients alive at 180 days demonstrated a statistically significant inverse association between medication count and readmission, with the model. On the other hand, logistic regression showed a significant positive association between problem count and readmission among survivors, indicating that each additional documented problem increased the odds

**Figure 4:**
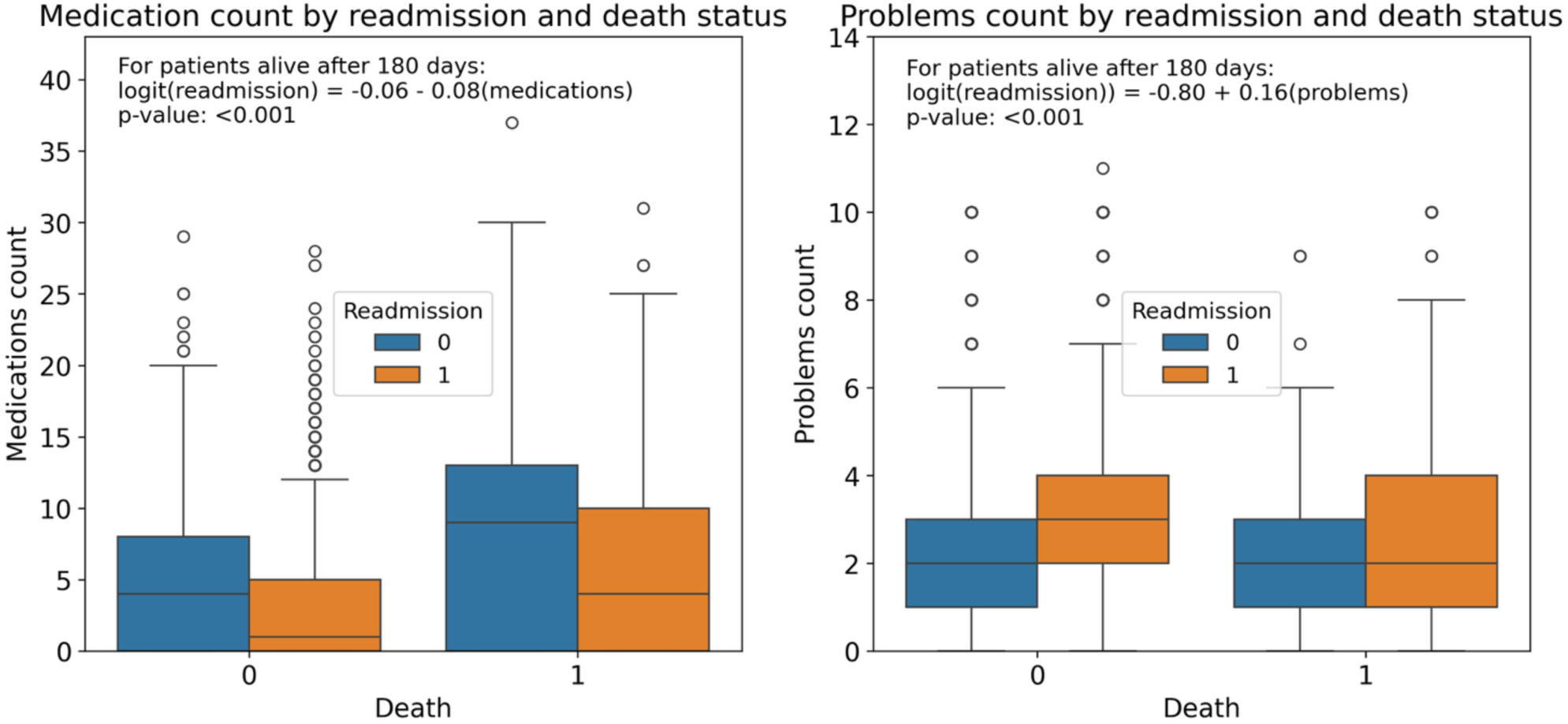
Boxplots showing the distribution of medication count (left panel) and problem count (right panel) according to 180-day mortality in the x-axis (Death = 0 or 1) and 180-day emergency readmission status (blue boxplots represent patients who were not readmitted, while the orange boxplots represent those who were readmitted). Results from a logistic regression analysis of readmission as a function of medication/problem count amongst survivors is also displayed.

Poor mobility and a high risk of falls were also associated with an increased probability of emergency readmission, whereas being married or in a de facto relationship appeared to have a protective effect. Mortality risk was primarily driven by higher urea levels, increased red blood cell distribution width (RDW-SD), lower systolic blood pressure, and older age. Reduced kidney function (lower eGFR), higher red cell distribution width (RDW-SD and RDW-CV), elevated urea levels, and older age were associated with an increased risk of both emergency readmission and death.

### 3.4 Visualising individual risk predictions

A useful approach to explaining individual-level predictions is to present each patient’s estimated risk together with the major feature contributions, expressed as changes in predicted probability derived from SHAP values. This enables clinicians to understand not only the overall predicted risk but also which variables most strongly increase or decrease the risk for a particular patient. The relative importance of individual predictors varied across patients, highlighting the heterogeneous drivers of adverse outcomes and the value of personalised risk visualisation.

To preserve patient privacy while illustrating individual predictions resembling clinically plausible scenarios, we generated representative synthetic patient profiles. Each synthetic profile was created by using clusters of 20 patients with similar risk prediction profiles. Within each cluster, continuous variables were summarised using the median, and categorical/binary variables using the most frequent category (mode) across the neighbourhood. Prediction models were used to estimate the risk of emergency readmission and death for these synthetic profiles. Figure 5 presents three synthetic patient profiles representing three clinically relevant risk scenarios: high risk of death (patient A), low risk of death but high risk of emergency readmission (patient B), and low risk of both outcomes (patient C).

**Figure 5:**
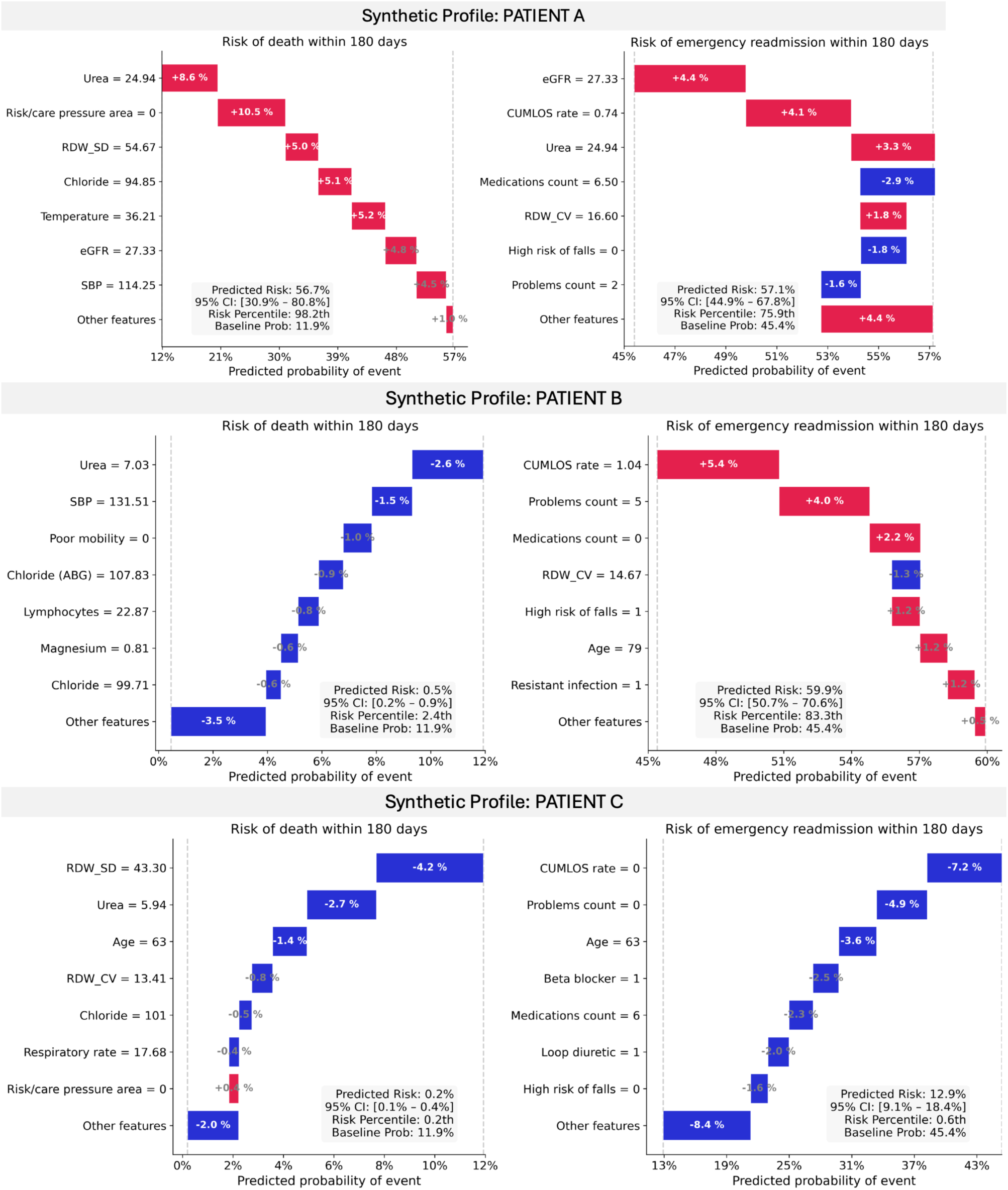
Waterfall plots illustrating how individual predictor variables (top 8) contribute to the models’ predicted probability of 180-day mortality (left) and emergency readmission (right) for three synthetic patients representative of: High risk of death (patient A), high risk of emergency readmission but low risk of death (patient B), and low risk of both readmission and death (patient C). The plot begins with the model’s baseline (average) predicted probability of event and sequentially shows the contribution of each feature to the final predicted risk. Features shown in red increase the predicted probability (positive contributions), whereas features shown in blue decrease the predicted probability (negative contributions). The length of each bar represents the magnitude of that variable’s effect on the prediction, with larger bars indicating a greater influence.

## 4. Discussion

In this study, we developed and evaluated clinically interpretable machine-learning models to predict emergency readmission and all-cause mortality within 180 days of hospital discharge among patients living with heart failure who were discharged alive and not recognised as approaching end of life. Using routinely collected electronic medical record (EMR) data linked with statewide administrative datasets, the models integrated structured clinical variables, longitudinal healthcare utilisation, and symptom information extracted from clinical notes. In addition to estimating predictive performance, we incorporated patient-level visualisation of risk drivers and prediction uncertainty to support clinically meaningful interpretation.

Our findings add to the growing evidence that adverse outcomes following hospitalisation for heart failure are common despite contemporary care. In our CardiacAI cohort, 45.2% of patients experienced at least one emergency readmission and 12.4% died within 180 days of discharge, despite not being reported to be at the end-of-life during the index admission. Within 30 days, 23.9% of patients had an emergency readmission and 2.6% had died. These findings are consistent with the Australian Institute of Health and Welfare’s national linked-data report (2025)^6^, which reported emergency readmission rates of 23% within 30 days and 64% within one year, alongside a 1-year mortality rate of 24% in a population characterised by high levels of frailty. Comparable 30-day and 1-year mortality estimates were described in an earlier systematic review and meta-analysis of Australian studies (Al-Omary et al. (2018)), which reported pooled all-cause readmission rates of 20% at 30 days and 56% at one year, with corresponding mortality rates of 8% at 30 days and 25% at one year. To our knowledge, the only other Australian study reporting 180-day readmission rates is a recent small retrospective cohort from a Melbourne hospital (Lammoza et al., 2025), which found a comparable readmission rate of 45.5%. The majority of emergency readmissions were attributable to recurrent HF, followed by atrial fibrillation, chest pain, pneumonia, acute kidney failure and chronic obstructive pulmonary disease. This pattern highlights the importance of integrated multidisciplinary follow-up rather than focusing exclusively on fluid management.

Our predictive algorithm for emergency readmission had a moderate performance (AUC=0.70 [0.66-0.73], Brier Score=0.22 [0.21-0.23]), and had reasonable calibration (calibration slope = 1.30) albeit with some under/over estimation of risk for high/low risk patients. Prediction of unplanned readmission after HF hospitalisation remains inherently challenging because readmission is influenced not only by disease severity but also by social support, treatment adherence, and health-system factors that are incompletely captured within electronic records. Few prior studies have specifically modelled unplanned readmission at 180 days following heart failure hospitalisation. A large (∼1.42 million patients) (Zheng et al 2022), retrospective cohort study using US administrative data found a modest AUC=0.59 for the prediction of all cause-readmission, likely due to the lack of clinical predictors. This agrees with findings from a review of traditional vs. machine learning (ML) models (Mortazavi et all 2016), which found that models of 180-day all-cause readmission had c-statistics ranging from 0.58 to 0.65, with the highest performing models being ML-based. By contrast, a small prospective highly curated study of 180-day readmission prediction in chronic heart failure (excluding patients with a planned readmission, missing data, dementia, sever liver and kidney dysfunction and cancer) (Gao et al 2021) reported a c-statistics 0.75 in an external dataset.

Previous healthcare utilisation emerged as the strongest predictor of emergency readmission, followed by the number of reported active clinical problems during the index admission, high falls risk, impaired mobility, reduced kidney function, elevated urea concentration and older age. These findings support the concept that recurrent hospitalisation reflects cumulative multimorbidity and persistent physiological vulnerability rather than isolated episodes of cardiac decompensation.

Patients discharged without key cardiovascular medications, particularly beta-blockers and loop diuretics, generally had a higher predicted risk of readmission. Several factors may explain this association. Patients not receiving these therapies may have had contraindications related to severe illness or haemodynamic instability, while incomplete capture of discharge medications because of imperfect medication reconciliation may also have contributed; also, very sick patients receiving medication may have died before readmission. Nevertheless, among patients who survived, absence of these core therapies remained associated with increased readmission risk, suggesting potentially modifiable opportunities for improving post-discharge care. These findings are consistent with Gao et al., who identified older age, previous acute heart failure, emergency admission and elevated blood urea nitrogen as independent predictors of readmission, whereas beta-blocker use was protective. Future analyses using time-to-event methods that explicitly account for competing risks will be needed to better characterise these associations.

The mortality prediction model had good performance (AUC=0.84 [0.80,0.88], Brier Score=0.08[0.07-0.09]) and was well calibrated (calibration slop = 1.01), although uncertainty around high risk patients was large due to the very small numbers. A systematic review and meta-analysis of risk stratification models for mortality in patients with HF (Siddiqi et al 2022) found c-statistics ranging from 0 .63 to 0.85. The best performing model was MARKER-HF (Adler et al 2018), a risk score including eight variables: Diastolic blood pressure, creatinine, blood urea nitrogen, haemoglobin, white blood cell count, platelets, albumin and red cell distribution width. Our study found elevated urea level, abnormal red blood cell distribution indices (RDW-SD, RDW-CV), older age, low systolic blood pressure, lymphopenia and hypochloremia were important predictors of mortality. RDW has consistently emerged as a strong prognostic marker in HF and may reflect multiple adverse biological processes including inflammation, malnutrition, renal dysfunction, impaired erythropoiesis, and physiological frailty.

Text-derived symptom variables, including nausea and poor mobility, also contributed meaningfully to mortality prediction. These features may capture symptom burden and functional impairment that are incompletely represented within structured EMR data, highlighting the additional prognostic value of information contained within clinical documentation.

An important strength of this study is the incorporation of patient-level visualisation of model predictions. Machine-learning algorithms are frequently criticised as “black boxes”, limiting clinician trust and adoption. By presenting SHAP-based feature contributions alongside predicted probabilities and measures of prediction uncertainty, benchmarked against the average risk within the cohort, clinicians can better understand the factors driving an individual patient’s predicted risk and assess whether the model’s reasoning aligns with clinical judgement. Such visualisations may also help identify potentially modifiable contributors to poor outcomes, including absence of guideline-directed therapy, falls risk, persistent renal dysfunction or recurrent congestion-related admissions. Although these models cannot establish causal relationships, they may assist clinicians in identifying patients who warrant closer review, targeted intervention and more intensive post-discharge follow-up.

### Limitations

Several limitations should be acknowledged. First, although the study included a comprehensive set of routinely collected clinical variables, the sample size was modest relative to the large number of candidate predictors. Model development using a larger cohort and external validation in other populations will be necessary to improve model performance and establish the generalisability of these models. Second, symptom extraction relied on keyword-based methods rather than contextual natural language processing. Although this pragmatic, privacy-preserving approach enabled extraction of clinically relevant information from free-text notes, it may not fully capture contextual nuance, temporal relationships, negation, or semantic ambiguity. Future work incorporating privacy-preserving large language models or clinical transformer architectures may improve identification of symptom burden, frailty, and functional status from clinical documentation.

Third, medication exposure at discharge may not accurately reflect actual treatment received after hospitalisation. Medication reconciliation was incomplete for a subset of admissions, and prescribed therapy does not necessarily indicate dispensing, adherence, or persistence following discharge. Also, the relationship between discharge medications, readmission, and mortality could be influenced by competing risks. Patients with advanced illness may die before experiencing readmission, while medications may be withheld because of contraindications or clinical instability rather than omission of evidence-based care. Future analyses using time-to-event methods with competing-risk models

## Conclusion

Routinely collected electronic medical record data, linked administrative datasets, and information extracted from clinical notes can be used to develop clinically interpretable machine-learning models for predicting adverse outcomes following heart failure hospitalisation. By combining risk prediction with transparent patient-level explanations of feature contributions and prediction uncertainty, these models have the potential to support clinical decision-making, identify patients at greatest risk, and highlight potentially modifiable factors that may benefit from closer review and targeted intervention.

### Ethics Approval

Approval was granted by the South Eastern Sydney Local Health District HREC Executive Committee (2019/ETH12625) & NSW Population Health Services Research EthicsCommittee (2020/ETH01614). CardiacAI operates under an opt-out model whereby posters are displayed in participating hospitals and information sheets are made available to patients on request.

## Data Availability

Data availability: The CardiacAI database and research activities are overseen by a Data Governance Committee. This committee is responsible for ensuring that all research is conducted within the ethical framework of the project and the data is kept secure and participant privacy is maintained. Information on data access requests can be found at caridacai.org.

https://www.cardiacai.org

## Acknowledgements

We acknowledge the use of CardiacAI data repository. We further acknowledge the contribution of the South Eastern Sydney (SESLHD) and Illawarra Shoalhaven (ISLHD) Local Health Districts, the NSW Ministry of Health for the use of NSW APDC and NSW RBDM, and the Centre for Health Record Linkage in facilitating access to the data used in this study.

## Data availability

The CardiacAI database and research activities are overseen by a Data Governance Committee. This committee is responsible for ensuring that all research is conducted within the ethical framework of the project and the data is kept secure and participant privacy is maintained. Information on data access requests can be found at caridacai.org.

