## Supplementary material for "Explainable, personalised prediction of emergency readmission and mortality following hospitalisation in patients with heart failure"

**Table 1**
Keywords used to extract concepts from text

- terms_EOLCare = ['palliative care', 'palliative treatment', 'palliative treatments', 'palliative therapy', 'palliative therapies', 'palliative (care team)', 'providing palliation', 'pal care', 'life-sustaining treatment', 'comfort care', 'supportive care', 'supportive cares', 'palliative supportive care', 'symptom management', 'symptoms management', 'withdrawal of life support', 'withdrawal of care', 'withdrawal of active care', 'withdrawal of life-sustaining treatment', 'terminal patient care', 'terminal care', 'dying care', 'caring for the dying', 'dying/death measures', 'end stage management', 'hospice care', 'end of life care']
- terms_pain = ['pain', 'painful']
- terms_pain_exclusion = ['no pain', 'nil pain', 'without pain', 'denies chest pain', 'no chest pain', 'nil chest pain', 'denies abdominal pain', 'no abdominal pain', 'nil abdominal pain', 'Pain score 0/10']
- terms_dyspnea = ['dyspnea', 'shortness of breath', 'SOB', 'short of breath', 'SOBOE', 'dyspnoeic', 'orthopnoea', 'tachypnoea', 'breathless', 'breathlessness']
- terms_dyspnea_exclusion = ['no dyspnea', 'no shortness of breath', 'no SOB', 'nil dyspnea', 'nil shortness of breath', 'nil SOB']
- terms_asthenia = ['asthenia', 'weakness', 'lack of energy', 'lack of strength', 'fatigue', 'exhaustion']
- terms_asthenia_exclusion = ['no asthenia', 'nil asthenia']
- terms_nausea = ['nausea', 'nauseous', 'urge to vomit', 'retching']
- terms_nausea_exclusion = ['no nausea', 'nil nausea', 'no nauseous', 'no vomiting or nausea', 'nil vomiting or nausea']
- terms_vomiting = ['vomiting', 'vomited', 'vomits', 'vomit', 'emesis', 'being sick', 'throwing up', 'throw up']
- terms_vomiting_exclusion = ['no vomiting', 'no nausea or vomiting', 'nil nausea or vomiting']
- terms_appetite = ['decrease in appetite', 'appetite decrease', 'appetite decreased', 'appetite decreasing', 'appetites decrease', 'decrease appetite', 'decreased food appetite', 'food appetite, decreased', 'loss of appetite', 'poor appetite', 'appetite poor', 'reduced appetite', 'appetite reducing', 'inappetence', 'hyporexia', 'appetite suppression', 'refusing food', 'hypophagia', 'anorexia', 'dysphagia']
- terms_appetite_exclusion = ['good appetite', 'normal appetite', 'no dysphagia', 'nil dysphagia', 'no hypophagia', 'nil hypophagia', 'no anorexia', 'nil anorexia', 'Poor appetite = No']
- terms_constipation = ['constipation', 'difficulty opening bowels', 'difficulty defecating', 'difficulty defaecating', 'difficulty passing motion', 'difficulty passing stool', 'difficult stool passage', 'constipated', 'constipate', 'constipating', 'costiveness', 'fecal retention', 'enema']
- terms_constipation_exclusion = ['no constipation', 'nil constipation', 'not constipated']
- terms_dry_mouth = ['sore mouth', 'mouth sore', 'xerostomia', 'xerostomias', 'asialia', 'asialias', 'hyposalivation', 'hyposalivations', 'mouth dryness', 'dryness mouth', 'salivary secretion decreased', 'decreased salivary secretion', 'dry mouth', 'mouth dry', 'drying mouth', 'absent salivary secretion', 'salivary secretion absent', 'aptyalism', 'saliva decreased', 'decreased saliva', 'oral dryness', 'aptyalia', 'hyposecretion of salivary gland', 'salivary gland

hyposecretion', 'salivary hyposecretion', 'reduced salivation', 'salivation decreased', 'decrease in salivation', 'hyposalivation', 'decreased salivary flow', 'mouth became dry', 'hypoactive salivary flow', 'inadequate salivary flow', 'salivary secretion disturbance']

- terms_dry_mouth_exclusion = ['no hyposalivation', 'nil hyposalivation', 'adequate salivary flow', 'normal salivation', 'no oral dryness', 'nil oral dryness']
- terms_drowsiness = ['somnolence', 'drowsiness', 'sleepiness', 'somnolent', 'hypersomnia', 'hypersomnolence', 'patient is drowsy']
- terms_drowsiness_exclusion = ['no somnolence', 'no drowsiness', 'nil somnolence', 'nil drowsiness', 'not drowsy', 'no hypersomnia', 'nil hypersomnia', 'fully alert']
- terms_poor_mobility = ['mobility poor', 'poor mobility', 'difficulty walking', 'impaired mobility', 'wheelchair dependent', 'wheelchair bound', 'limited mobibility', 'bed bound', 'bedbound', 'poor joint stability', 'mobilises with 4WW', 'mobilises with 4WWD', 'mobilises with walker', 'mobilises with frame', 'mobilises with walking stick', 'mobilises with aid', 'mobilises with wheelchair', 'ambulant with 4WW', 'ambulant with 4WWD', 'ambulant with walker', 'ambulant with frame', 'ambulant with walking stick', 'ambulant with aid', 'ambulant with wheelchair', 'requires 4WW', 'requires 4WWD', 'requires walker', 'requires frame', 'requires walking stick', 'requires mobility aid', 'with 4WW', 'with 4WWD', 'with walker', 'with frame', 'with walking stick', 'with mobility aid', 'requires wheelchair', 'uses 4WW', 'uses 4WWD', 'uses walker', 'uses frame', 'uses walking stick', 'uses walking stick', 'uses mobility aid', 'uses wheelchair', 'walks with help']
- terms_poor_mobility_exclusion = ['normal mobility', 'good mobility', 'mobilising independently with nil aid']
- terms_depression = ['depressed mood', 'mood depressed', 'depression', 'depressed', 'feeling low', 'low mood', 'melancholic', 'miserable', 'sad', 'morose mood', 'morosity', 'melancholy', 'decreased mood', 'feeling blue', 'mood depression', 'depression mood', 'feeling down', 'depressive disorder', 'depressive symptoms', 'anhedonia', 'persistent sadness']
- terms_depression_exclusion = ['no depression', 'nil depression', 'not depressed', 'no depressive', 'nil depressive', 'ST depression', 'no depressive symptoms', 'no anhedonia', 'nil depressive symptoms', 'nil anhedonia']
- terms_fraility = ['mildly frail', 'moderately frail', 'severely frail', 'is frail', 'mild frailty', 'moderate frailty', 'severe frailty', 'shows frailty', 'frailty syndrome', 'Clinical Frailty score - 5', 'Clinical Frailty score - 6', 'Clinical Frailty score - 7', 'Clinical Frailty score - 8', 'Clinical Frailty score - 9', 'CFS - 5', 'CFS - 6', 'CFS - 7', 'CFS - 8', 'CFS - 9', 'Edmonton frail score = 8', 'Edmonton frail score = 9', 'Edmonton frail score = 10', 'Edmonton frail score = 11', 'Edmonton frail score = 12', 'Edmonton frail score = 13', 'Edmonton frail score = 14', 'Edmonton frail score = 15', 'Edmonton frail score = 16', 'Edmonton frail score = 17', 'Edmonton frail score = 18', 'Essential Frailty Score - score = 3', 'Essential Frailty Score - score = 4', 'Essential Frailty Score - score = 5', 'EFS - score = 3', 'EFS - score = 4', 'EFS - score = 5', 'Frailty Screening Tool Score : 3', 'Frailty Screening Tool Score : 4', 'Frailty Screening Tool Score : 5']
- terms_fraility_exclusion = ['not frail', 'no frailty', 'nil frailty']
- terms_deterioration = ['deterioration', 'deteriorates', 'deteriorating', 'condition worsening', 'decline in condition', 'decompensation', 'worsening of condition', 'functional decline', 'continue to decline', 'continue to deteriorate', 'decreased function']
- terms_deterioration_exclusion = ['no evidence of decompensation', 'no evidence of deterioration']
- terms_adl = ['reduced activities of daily living', 'reduced ADL', 'dependent for ADL', 'requires assistant with ADL', 'requires assistant with activities of daily living', 'dependent for activities of daily living', 'reduced activities of daily living', 'increased care needs']
- terms_adl_exclusion = ['independent ADL']

- terms_HFpEF = ['HFpEF', 'preserved ejection fraction', 'preserved EF', 'preserved LV', 'preserved LVEF', 'normal LV', 'normal LVEF']
- terms_HFpEF_exclusion = ['not preserved ejection fraction', 'not preserved EF', 'not preserved LV', 'not preserved LVEF', 'HFrEF', 'reduced ejection fraction', 'reduced EF', 'reduced LV', 'reduced LVEF', 'low EF', 'low LVEF']
- terms_HFrEF = ['HFrEF', 'reduced ejection fraction', 'reduced EF', 'reduced LV', 'reduced LVEF', 'low EF', 'low LVEF', 'EF <40%', 'EF <=40%', 'EF 40%', 'EF 39%', 'EF 38%', 'EF 37%', 'EF 36%', 'EF 35%', 'EF 34%', 'EF 33%', 'EF 32%', 'EF 31%', 'EF 30%', 'EF 29%', 'EF 28%', 'EF 27%', 'EF 26%', 'EF 25%', 'EF 24%', 'EF 23%', 'EF 22%', 'EF 21%', 'EF 20%', 'EF 19%', 'EF 18%', 'EF 17%', 'EF 16%', 'EF 15%', 'EF 14%', 'EF 13%', 'EF 12%', 'EF 11%', 'EF 10%', 'EF 9%', 'EF 8%', 'EF 7%', 'EF 6%', 'EF=40%', 'EF=39%', 'EF=38%', 'EF=37%', 'EF=36%', 'EF=35%', 'EF=34%', 'EF=33%', 'EF=32%', 'EF=31%', 'EF=30%', 'EF=29%', 'EF=28%', 'EF=27%', 'EF=26%', 'EF=25%', 'EF=24%', 'EF=23%', 'EF=22%', 'EF=21%', 'EF=20%', 'EF=19%', 'EF=18%', 'EF=17%', 'EF=16%', 'EF=15%', 'EF=14%', 'EF=13%', 'EF=12%', 'EF=11%', 'EF=10%', 'EF=9%', 'EF=8%', 'EF=7%', 'EF=6%', 'LVEF <40%', 'LVEF <=40%', 'LVEF 40%', 'LVEF 39%', 'LVEF 38%', 'LVEF 37%', 'LVEF 36%', 'LVEF 35%', 'LVEF 34%', 'LVEF 33%', 'LVEF 32%', 'LVEF 31%', 'LVEF 30%', 'LVEF 29%', 'LVEF 28%', 'LVEF 27%', 'LVEF 26%', 'LVEF 25%', 'LVEF 24%', 'LVEF 23%', 'LVEF 22%', 'LVEF 21%', 'LVEF 20%', 'LVEF 19%', 'LVEF 18%', 'LVEF 17%', 'LVEF 16%', 'LVEF 15%', 'LVEF 14%', 'LVEF 13%', 'LVEF 12%', 'LVEF 11%', 'LVEF 10%', 'LVEF 9%', 'LVEF 8%', 'LVEF 7%', 'LVEF 6%', 'LVEF=40%', 'LVEF=39%', 'LVEF=38%', 'LVEF=37%', 'LVEF=36%', 'LVEF=35%', 'LVEF=34%', 'LVEF=33%', 'LVEF=32%', 'LVEF=31%', 'LVEF=30%', 'LVEF=29%', 'LVEF=28%', 'LVEF=27%', 'LVEF=26%', 'LVEF=25%', 'LVEF=24%', 'LVEF=23%', 'LVEF=22%', 'LVEF=21%', 'LVEF=20%', 'LVEF=19%', 'LVEF=18%', 'LVEF=17%', 'LVEF=16%', 'LVEF=15%', 'LVEF=14%', 'LVEF=13%', 'LVEF=12%', 'LVEF=11%', 'LVEF=10%', 'LVEF=9%', 'LVEF=8%', 'LVEF=7%', 'LVEF=6%']
- terms_HFrEF_exclusion = ['no HFrEF', 'not reduced ejection fraction', 'not reduced EF', 'not reduced LV', 'not reduced LVEF', 'nil HFrEF', 'nil reduced ejection fraction', 'nil reduced EF', 'nil reduced LV', 'nil reduced LVEF', 'normal EF', 'normal LVEF']

**Table 2**
Medication groupings

| Medication name | Primary_group |
|---|---|
| mirtazapine | CNS: Antipsychotic |
| quetiapine | CNS: Antipsychotic |
| olanzapine | CNS: Antipsychotic |
| clozapine | CNS: Antipsychotic |
| risperidone | CNS: Antipsychotic |
| haloperidol | CNS: Antipsychotic |
| lurasidone | CNS: Antipsychotic |
| oxazepam | CNS: Benzodiazepine |
| temazepam | CNS: Benzodiazepine |
| diazepam | CNS: Benzodiazepine |
| bromazepam | CNS: Benzodiazepine |
| alprazolam | CNS: Benzodiazepine |
| nitrazepam | CNS: Benzodiazepine |

| | |
|---|---|
| clonazepam | CNS: Benzodiazepine |
| lorazepam | CNS: Benzodiazepine |
| flunitrazepam | CNS: Benzodiazepine |
| zolpidem | CNS: Hypnotic (Z-drug) |
| pregabalin | CNS: Mood stabiliser/antiepileptic |
| levetiracetam | CNS: Mood stabiliser/antiepileptic |
| gabapentin | CNS: Mood stabiliser/antiepileptic |
| valproate | CNS: Mood stabiliser/antiepileptic |
| carbamazepine | CNS: Mood stabiliser/antiepileptic |
| lamotrigine | CNS: Mood stabiliser/antiepileptic |
| sertraline | CNS: SSRI/SNRI antidepressant |
| duloxetine | CNS: SSRI/SNRI antidepressant |
| venlafaxine | CNS: SSRI/SNRI antidepressant |
| citalopram | CNS: SSRI/SNRI antidepressant |
| escitalopram | CNS: SSRI/SNRI antidepressant |
| paroxetine | CNS: SSRI/SNRI antidepressant |
| fluoxetine | CNS: SSRI/SNRI antidepressant |
| amitriptyline | CNS: TCA antidepressant |
| imipramine | CNS: TCA antidepressant |
| nortriptyline | CNS: TCA antidepressant |
| perindopril | CV: ACE inhibitor |
| ramipril | CV: ACE inhibitor |
| amlodipine-perindopril | CV: ACE inhibitor |
| quinapril | CV: ACE inhibitor |
| trandolapril | CV: ACE inhibitor |
| enalapril | CV: ACE inhibitor |
| lisinopril | CV: ACE inhibitor |
| enalapril-lercanidipine | CV: ACE inhibitor |
| indapamide-perindopril | CV: ACE inhibitor |
| fosinopril | CV: ACE inhibitor |
| fosinopril-hydrochlorothiazide | CV: ACE inhibitor |
| dutasteride-tamsulosin | CV: Alpha blocker |
| tamsulosin | CV: Alpha blocker |
| terazosin | CV: Alpha blocker |
| sotalol | CV: Antiarrhythmic |
| amiodarone | CV: Antiarrhythmic |
| flecainide | CV: Antiarrhythmic |
| apixaban | CV: Anticoagulant (DOAC) |
| dabigatran | CV: Anticoagulant (DOAC) |
| rivaroxaban | CV: Anticoagulant (DOAC) |
| enoxaparin | CV: Anticoagulant (warfarin/heparin) |
| warfarin | CV: Anticoagulant (warfarin/heparin) |
| heparin | CV: Anticoagulant (warfarin/heparin) |

| | |
|---|---|
| aspirin | CV: Antiplatelet |
| clopidogrel | CV: Antiplatelet |
| aspirin-clopidogrel | CV: Antiplatelet |
| ticagrelor | CV: Antiplatelet |
| prasugrel | CV: Antiplatelet |
| aspirin-dipyridamole | CV: Antiplatelet |
| telmisartan | CV: ARB |
| valsartan | CV: ARB |
| hydrochlorothiazide-olmesartan | CV: ARB |
| candesartan-hydrochlorothiazide | CV: ARB |
| sacubitril-valsartan | CV: ARB |
| irbesartan | CV: ARB |
| candesartan | CV: ARB |
| hydrochlorothiazide-irbesartan | CV: ARB |
| losartan | CV: ARB |
| amlodipine-olmesartan | CV: ARB |
| olmesartan | CV: ARB |
| hydrochlorothiazide-telmisartan | CV: ARB |
| amlodipine/hydrochlorothiazide/valsartan | CV: ARB |
| amlodipine-telmisartan | CV: ARB |
| amlodipine-valsartan | CV: ARB |
| amlodipine/hydrochlorothiazide/olmesartan | CV: ARB |
| hydrochlorothiazide-valsartan | CV: ARB |
| bisoprolol | CV: Beta blocker |
| metoprolol | CV: Beta blocker |
| nebivolol | CV: Beta blocker |
| dorzolamide-timolol ophthalmic | CV: Beta blocker |
| betaxolol ophthalmic | CV: Beta blocker |
| latanoprost-timolol ophthalmic | CV: Beta blocker |
| propranolol | CV: Beta blocker |
| carvedilol | CV: Beta blocker |
| atenolol | CV: Beta blocker |
| bimatoprost-timolol ophthalmic | CV: Beta blocker |
| timolol ophthalmic | CV: Beta blocker |
| labetalol | CV: Beta blocker |
| brimonidine-timolol ophthalmic | CV: Beta blocker |
| timolol-travoprost ophthalmic | CV: Beta blocker |
| brinzolamide-timolol ophthalmic | CV: Beta blocker |
| amlodipine | CV: Calcium channel blocker |
| lercanidipine | CV: Calcium channel blocker |
| diltiazem | CV: Calcium channel blocker |
| verapamil | CV: Calcium channel blocker |
| amlodipine-atorvastatin | CV: Calcium channel blocker |

| | |
|---|---|
| nifedipine | CV: Calcium channel blocker |
| felodipine | CV: Calcium channel blocker |
| digoxin | CV: Digoxin |
| furosemide | CV: Loop diuretic |
| bumetanide | CV: Loop diuretic |
| spironolactone | CV: MRA |
| eplerenone | CV: MRA |
| glyceryl trinitrate | CV: Nitrates/antianginal |
| isosorbide mononitrate | CV: Nitrates/antianginal |
| isosorbide dinitrate | CV: Nitrates/antianginal |
| methyldopa | CV: Other antihypertensive |
| hydralazine | CV: Other antihypertensive |
| clonidine | CV: Other antihypertensive |
| minoxidil | CV: Other antihypertensive |
| ezetimibe | CV: Other lipid-lowering |
| fenofibrate | CV: Other lipid-lowering |
| gemfibrozil | CV: Other lipid-lowering |
| sildenafil | CV: Pulmonary hypertension |
| tadalafil | CV: Pulmonary hypertension |
| macitentan | CV: Pulmonary hypertension |
| ambrisentan | CV: Pulmonary hypertension |
| riociguat | CV: Pulmonary hypertension |
| rosuvastatin | CV: Statin |
| atorvastatin | CV: Statin |
| simvastatin | CV: Statin |
| ezetimibe-simvastatin | CV: Statin |
| nystatin | CV: Statin |
| pravastatin | CV: Statin |
| atorvastatin-ezetimibe | CV: Statin |
| ezetimibe-rosuvastatin | CV: Statin |
| gramicidin/neomycin/nystatin/triamcinolone otic | CV: Statin |
| indapamide | CV: Thiazide diuretic |
| amiloride-hydrochlorothiazide | CV: Thiazide diuretic |
| hydrochlorothiazide | CV: Thiazide diuretic |
| prednisolone | Endo: Corticosteroid systemic |
| prednisone | Endo: Corticosteroid systemic |
| phenylephrine-prednisolone ophthalmic | Endo: Corticosteroid systemic |
| clotrimazole-hydrocortisone topical | Endo: Corticosteroid systemic |
| dexamethasone ophthalmic | Endo: Corticosteroid systemic |
| cinchocaine-hydrocortisone topical | Endo: Corticosteroid systemic |
| dexamethasone | Endo: Corticosteroid systemic |
| prednisolone ophthalmic | Endo: Corticosteroid systemic |
| methylprednisolone topical | Endo: Corticosteroid systemic |

| | |
|---|---|
| hydrocortisone | Endo: Corticosteroid systemic |
| fludrocortisone | Endo: Corticosteroid systemic |
| hydrocortisone topical | Endo: Corticosteroid systemic |
| linagliptin | Endo: DPP-4 inhibitor |
| vildagliptin | Endo: DPP-4 inhibitor |
| sitagliptin | Endo: DPP-4 inhibitor |
| ertugliflozin-sitagliptin | Endo: DPP-4 inhibitor |
| saxagliptin | Endo: DPP-4 inhibitor |
| empagliflozin-linagliptin | Endo: DPP-4 inhibitor |
| exenatide | Endo: GLP-1 RA |
| dulaglutide | Endo: GLP-1 RA |
| liraglutide | Endo: GLP-1 RA |
| insulin aspart-insulin aspart protamine | Endo: Insulin |
| insulin glargine | Endo: Insulin |
| insulin aspart | Endo: Insulin |
| insulin glulisine | Endo: Insulin |
| insulin detemir | Endo: Insulin |
| insulin isophane-insulin neutral | Endo: Insulin |
| insulin neutral | Endo: Insulin |
| insulin isophane | Endo: Insulin |
| insulin lispro | Endo: Insulin |
| insulin aspart-insulin degludec | Endo: Insulin |
| insulin lispro-insulin lispro protamine | Endo: Insulin |
| metformin | Endo: Metformin |
| linagliptin-metformin | Endo: Metformin |
| metformin-sitagliptin | Endo: Metformin |
| empagliflozin-metformin | Endo: Metformin |
| glibenclamide-metformin | Endo: Metformin |
| metformin-vildagliptin | Endo: Metformin |
| dapagliflozin-metformin | Endo: Metformin |
| metformin-saxagliptin | Endo: Metformin |
| denosumab | Endo: Osteoporosis |
| alendronate-colecalciferol | Endo: Osteoporosis |
| risedronate sodium | Endo: Osteoporosis |
| raloxifene | Endo: Osteoporosis |
| calcium carbonate-risedronate sodium | Endo: Osteoporosis |
| alendronate | Endo: Osteoporosis |
| alendronate/calcium co3/colecalciferol | Endo: Osteoporosis |
| testosterone | Endo: Sex hormones |
| estradiol | Endo: Sex hormones |
| dydrogesterone-estradiol | Endo: Sex hormones |
| estradiol topical | Endo: Sex hormones |
| estradiol-norethisterone | Endo: Sex hormones |

| | |
|---|---|
| progesterone | Endo: Sex hormones |
| dapagliflozin | Endo: SGLT2 inhibitor |
| empagliflozin | Endo: SGLT2 inhibitor |
| gliclazide | Endo: Sulfonylurea |
| glimepiride | Endo: Sulfonylurea |
| glipizide | Endo: Sulfonylurea |
| glibenclamide | Endo: Sulfonylurea |
| levothyroxine sodium | Endo: Thyroid hormone/antithyroid |
| carbimazole | Endo: Thyroid hormone/antithyroid |
| propylthiouracil | Endo: Thyroid hormone/antithyroid |
| pioglitazone | Endo: TZD |
| loperamide | GI: Antidiarrheal |
| atropine-diphenoxylate | GI: Antidiarrheal |
| ondansetron | GI: Antiemetic |
| promethazine | GI: Antiemetic |
| metoclopramide | GI: Antiemetic |
| prochlorperazine | GI: Antiemetic |
| ranitidine | GI: H2 blocker |
| nizatidine | GI: H2 blocker |
| famotidine | GI: H2 blocker |
| sulfasalazine | GI: IBD |
| ustekinumab | GI: IBD |
| aluminium hydroxide/magnesium hydroxide/simethicone | GI: Laxative |
| docusate | GI: Laxative |
| docusate-senna | GI: Laxative |
| methoxy polyethylene glycol-epoetin beta | GI: Laxative |
| lactulose | GI: Laxative |
| senna | GI: Laxative |
| psyllium | GI: Laxative |
| bisacodyl | GI: Laxative |
| pantoprazole | GI: PPI |
| esomeprazole | GI: PPI |
| rabeprazole | GI: PPI |
| omeprazole | GI: PPI |
| aripiprazole | GI: PPI |
| lansoprazole | GI: PPI |
| ascorbic acid-ferrous sulfate | Heme: ESA/iron |
| darbepoetin alfa | Heme: ESA/iron |
| ferrous fumarate | Heme: ESA/iron |
| ferrous fumarate-folic acid | Heme: ESA/iron |
| ferrous sulfate | Heme: ESA/iron |
| ferrous sulfate-folic acid | Heme: ESA/iron |
| iron polymaltose | Heme: ESA/iron |

| | |
|---|---|
| multivitamin with iron | Heme: ESA/iron |
| epoetin beta | Heme: ESA/iron |
| gentamicin | ID: Aminoglycoside |
| clotrimazole topical | ID: Antifungal |
| itraconazole | ID: Antifungal |
| miconazole | ID: Antifungal |
| amphotericin b | ID: Antifungal |
| fluconazole | ID: Antifungal |
| bifonazole topical | ID: Antifungal |
| ketoconazole topical | ID: Antifungal |
| emtricitabine-tenofovir | ID: Antiviral |
| tenofovir | ID: Antiviral |
| oseltamivir | ID: Antiviral |
| ledipasvir-sofosbuvir | ID: Antiviral |
| bictegravir/emtricitabine/tenofovir | ID: Antiviral |
| cefalexin | ID: Cephalosporin |
| cefuroxime | ID: Cephalosporin |
| cefaclor | ID: Cephalosporin |
| ceftriaxone | ID: Cephalosporin |
| cefazolin | ID: Cephalosporin |
| ciprofloxacin | ID: Fluoroquinolone |
| ciprofloxacin otic | ID: Fluoroquinolone |
| norfloxacin | ID: Fluoroquinolone |
| ofloxacin ophthalmic | ID: Fluoroquinolone |
| moxifloxacin | ID: Fluoroquinolone |
| ciprofloxacin ophthalmic | ID: Fluoroquinolone |
| clindamycin | ID: Lincosamide |
| azithromycin | ID: Macrolide |
| roxithromycin | ID: Macrolide |
| clarithromycin | ID: Macrolide |
| metronidazole topical | ID: Nitroimidazole |
| metronidazole | ID: Nitroimidazole |
| amoxicillin-clavulanate | ID: Penicillin/beta-lactam |
| amoxicillin | ID: Penicillin/beta-lactam |
| flucloxacillin | ID: Penicillin/beta-lactam |
| benzylpenicillin sodium | ID: Penicillin/beta-lactam |
| benzathine benzylpenicillin | ID: Penicillin/beta-lactam |
| ampicillin | ID: Penicillin/beta-lactam |
| sulfamethoxazole-trimethoprim | ID: Sulfonamide/trimethoprim |
| trimethoprim | ID: Sulfonamide/trimethoprim |
| doxycycline | ID: Tetracycline |
| methotrexate | Immuno: DMARD/biologic |
| leflunomide | Immuno: DMARD/biologic |

| | |
|---|---|
| ranibizumab ophthalmic | Immuno: DMARD/biologic |
| baricitinib | Immuno: DMARD/biologic |
| tocilizumab | Immuno: DMARD/biologic |
| macrogol 3350 with electrolytes | Other |
| multivitamin with minerals | Other |
| latanoprost ophthalmic | Other |
| estriol topical | Other |
| allopurinol | Other |
| betamethasone topical | Other |
| colecalciferol | Other |
| sodium polystyrene sulfonate | Other |
| calamine topical | Other |
| hydroxocobalamin | Other |
| prazosin | Other |
| bromhexine | Other |
| multivitamin | Other |
| omega-3 fatty acids | Other |
| ocular lubricant | Other |
| melatonin | Other |
| monobasic sodium phosphate | Other |
| zinc sulfate | Other |
| moxonidine | Other |
| hydroxychloroquine | Other |
| atropine ophthalmic | Other |
| terbutaline | Other |
| tapentadol | Other |
| emollients topical | Other |
| ascorbic acid | Other |
| ascorbic acid-cranberry | Other |
| methenamine hippurate | Other |
| colchicine | Other |
| hydroxycarbamide | Other |
| nicorandil | Other |
| donepezil | Other |
| doxepin | Other |
| methyl salicylate topical | Other |
| sodium chloride nasal | Other |
| frangula-sterculia | Other |
| zinc oxide topical | Other |
| goserelin | Other |
| triamcinolone topical | Other |
| venetoclax | Other |
| cabergoline | Other |

| | |
|---|---|
| quinine | Other |
| aciclovir ophthalmic | Other |
| domperidone | Other |
| indometacin | Other |
| sodium chloride | Other |
| desvenlafaxine | Other |
| perhexiline | Other |
| glucosamine | Other |
| sodium hyaluronate ophthalmic | Other |
| rivastigmine | Other |
| citric acid/mg oxide/sodium picosulfate | Other |
| baclofen | Other |
| cinchocaine-zinc oxide topical | Other |
| lipase/amylase/protease | Other |
| glycopyrronium | Other |
| balsalazide | Other |
| betahistine | Other |
| carbidopa/entacapone/levodopa | Other |
| selegiline | Other |
| pramipexole | Other |
| carbidopa-levodopa | Other |
| aluminium hydroxide | Other |
| acetazolamide | Other |
| calcitriol | Other |
| sodium fusidate | Other |
| balsam peru/benzyl benzoate/zinc oxide topical | Other |
| theophylline | Other |
| imatinib | Other |
| alpha-tocopherol | Other |
| amisulpride | Other |
| cortisone | Other |
| valaciclovir | Other |
| galantamine | Other |
| brimonidine ophthalmic | Other |
| cinacalcet | Other |
| tetryzoline ophthalmic | Other |
| sodium cromoglycate | Other |
| letrozole | Other |
| acitretin | Other |
| etacrynic acid | Other |
| pyridoxine | Other |
| cyproterone | Other |
| etanercept | Other |

| | |
|---|---|
| benserazide-levodopa | Other |
| bimatoprost ophthalmic | Other |
| capsaicin topical | Other |
| agomelatine | Other |
| ubidecarenone | Other |
| silver sulfadiazine topical | Other |
| lidocaine topical | Other |
| mianserin | Other |
| chloramphenicol ophthalmic | Other |
| eucalyptus/menthol/methyl salicylate topical | Other |
| glucagon | Other |
| povidone iodine topical | Other |
| wheat dextrin | Other |
| safinamide | Other |
| rotigotine | Other |
| simethicone | Other |
| phenytoin | Other |
| azathioprine | Other |
| glycerol | Other |
| colestyramine | Other |
| rasagiline | Other |
| travoprost ophthalmic | Other |
| enzalutamide | Other |
| al hydroxide/mg hydroxide/mg trisilicate | Other |
| lidocaine -zinc oxide topical | Other |
| beclometasone | Other |
| primidone | Other |
| triamcinolone | Other |
| nedocromil | Other |
| heparinised saline | Other |
| mesalazine | Other |
| saliva substitutes | Other |
| terbinafine | Other |
| hydromorphone | Other |
| brimonidine-brinzolamide ophthalmic | Other |
| misoprostol | Other |
| prucalopride | Other |
| anastrozole | Other |
| vortioxetine | Other |
| calcipotriol topical | Other |
| mexiletine | Other |
| fluvoxamine | Other |
| fluorometholone ophthalmic | Other |

| | |
|---|---|
| fosfomycin | Other |
| pentoxifylline | Other |
| pilocarpine ophthalmic | Other |
| acarbose | Other |
| sumatriptan | Other |
| brinzolamide ophthalmic | Other |
| aflibercept ophthalmic | Other |
| medroxyprogesterone | Other |
| nitrofurantoin | Other |
| heparinoids topical | Other |
| choline salicylate topical | Other |
| chlortalidone | Other |
| permethrin topical | Other |
| ciclosporin | Other |
| dosulepin | Other |
| doxylamine | Other |
| sucralfate | Other |
| macrogol 3350 | Other |
| midodrine | Other |
| ferric carboxymaltose | Other |
| reboxetine | Other |
| paliperidone | Other |
| rifampicin | Other |
| dimethyl fumarate | Other |
| betamethasone-calcipotriol topical | Other |
| bicalutamide | Other |
| octreotide | Other |
| cyclizine | Other |
| olsalazine | Other |
| selenium | Other |
| dorzolamide ophthalmic | Other |
| lithium | Other |
| bifidobacterium-lactobacillus | Other |
| ibrutinib | Other |
| phytomenadione | Other |
| trihexyphenidyl | Other |
| ursodeoxycholic acid | Other |
| memantine | Other |
| pirfenidone | Other |
| mupirocin topical | Other |
| drospirenone-ethinylestradiol | Other |
| anagrelide | Other |
| leuprorelin | Other |

| | |
|---|---|
| mebeverine | Other |
| electrolyte replacement solutions oral | Other |
| tamoxifen | Other |
| pamidronate disodium | Other |
| lenalidomide | Other |
| ammonium bicarbonate-senega | Other |
| clomipramine | Other |
| disopyramide | Other |
| pyridostigmine | Other |
| naltrexone | Other |
| tranylcypromine | Other |
| pristinamycin | Other |
| urea topical | Other |
| lamivudine | Other |
| tramazoline nasal | Other |
| fluorouracil topical | Other |
| beclometasone nasal | Other |
| probenecid | Other |
| pizotifen | Other |
| paraffin liquid | Other |
| rifaximin | Other |
| carbamide peroxide otic | Other |
| entecavir | Other |
| mycophenolic acid | Other |
| cyclophosphamide | Other |
| ruxolitinib | Other |
| paracetamol | Pain: Acetaminophen/paracetamol |
| meloxicam | Pain: NSAID |
| diclofenac topical | Pain: NSAID |
| celecoxib | Pain: NSAID |
| ibuprofen | Pain: NSAID |
| diclofenac | Pain: NSAID |
| ketorolac ophthalmic | Pain: NSAID |
| ibuprofen-paracetamol | Pain: NSAID |
| naproxen | Pain: NSAID |
| oxycodone | Pain: Opioid |
| naloxone-oxycodone | Pain: Opioid |
| buprenorphine | Pain: Opioid |
| morphine | Pain: Opioid |
| tramadol | Pain: Opioid |
| codeine-paracetamol | Pain: Opioid |
| buprenorphine-naloxone | Pain: Opioid |
| methadone | Pain: Opioid |

| | |
|---|---|
| codeine | Pain: Opioid |
| codeine/doxylamine/paracetamol | Pain: Opioid |
| fentanyl | Pain: Opioid |
| sevelamer | Renal: Phosphate binder |
| aclidinium | Resp: Anticholinergic bronchodilator |
| ipratropium | Resp: Anticholinergic bronchodilator |
| umeclidinium | Resp: Anticholinergic bronchodilator |
| tiotropium | Resp: Anticholinergic bronchodilator |
| umeclidinium-vilanterol | Resp: Anticholinergic bronchodilator |
| fluticasone/umeclidinium/vilanterol | Resp: Anticholinergic bronchodilator |
| ipratropium nasal | Resp: Anticholinergic bronchodilator |
| ivabradine | Resp: Antihistamine |
| loratadine | Resp: Antihistamine |
| fexofenadine | Resp: Antihistamine |
| cetirizine | Resp: Antihistamine |
| cyproheptadine | Resp: Antihistamine |
| olopatadine ophthalmic | Resp: Antihistamine |
| amantadine | Resp: Antihistamine |
| salbutamol | Resp: Beta-agonist bronchodilator |
| budesonide-formoterol | Resp: Beta-agonist bronchodilator |
| fluticasone-formoterol | Resp: Beta-agonist bronchodilator |
| fluticasone-salmeterol | Resp: Beta-agonist bronchodilator |
| olodaterol-tiotropium | Resp: Beta-agonist bronchodilator |
| glycopyrronium -indacaterol | Resp: Beta-agonist bronchodilator |
| indacaterol | Resp: Beta-agonist bronchodilator |
| aclidinium-formoterol | Resp: Beta-agonist bronchodilator |
| salmeterol | Resp: Beta-agonist bronchodilator |
| pseudoephedrine | Resp: Decongestant |
| oxymetazoline nasal | Resp: Decongestant |
| fluticasone-vilanterol | Resp: Inhaled corticosteroid |
| fluticasone | Resp: Inhaled corticosteroid |
| mometasone nasal | Resp: Inhaled corticosteroid |
| budesonide nasal | Resp: Inhaled corticosteroid |
| budesonide | Resp: Inhaled corticosteroid |
| azelastine-fluticasone nasal | Resp: Inhaled corticosteroid |
| ciclesonide | Resp: Inhaled corticosteroid |
| mometasone topical | Resp: Inhaled corticosteroid |
| fluticasone nasal | Resp: Inhaled corticosteroid |
| montelukast | Resp: Leukotriene modifier |
| nicotine | Resp: Smoking cessation |
| varenicline | Resp: Smoking cessation |
| magnesium aspartate | Supp: Electrolyte/mineral |
| sodium bicarbonate | Supp: Electrolyte/mineral |

| | |
|---|---|
| calcium carbonate | Supp: Electrolyte/mineral |
| alginic acid/calcium carbonate/sodium bicarbonate | Supp: Electrolyte/mineral |
| potassium chloride | Supp: Electrolyte/mineral |
| alginic acid/calcium carbonate/potassium bicarbonate | Supp: Electrolyte/mineral |
| citric acid/sodium bicarbonate/sodium citrate/tartaric acid | Supp: Electrolyte/mineral |
| calcium-colecalciferol | Supp: Electrolyte/mineral |
| sodium bicarbonate topical | Supp: Electrolyte/mineral |
| potassium bicarbonate/potassium carbonate/potassium chloride | Supp: Electrolyte/mineral |
| calcium citrate | Supp: Electrolyte/mineral |
| magnesium sulfate | Supp: Electrolyte/mineral |
| calcium polystyrene sulfonate | Supp: Electrolyte/mineral |
| citric acid/sodium bicarbonate/tartaric acid | Supp: Electrolyte/mineral |
| folic acid | Supp: Vitamin |
| thiamine | Supp: Vitamin |
| cyanocobalamin | Supp: Vitamin |
| vitamin a ophthalmic | Supp: Vitamin |
| tacrolimus | Transplant: Calcineurin/mTOR |
| mycophenolate mofetil | Transplant: Calcineurin/mTOR |
| dutasteride | Uro: BPH |
| finasteride | Uro: BPH |
| oxybutynin | Uro: Overactive bladder |
| mirabegron | Uro: Overactive bladder |
| solifenacin | Uro: Overactive bladder |
| influenza virus vaccine inactivated | Vaccine/Immunisation |
| influenza virus vaccine h1n1 inactivated | Vaccine/Immunisation |
| pneumococcal 23-valent polysaccharide vaccine | Vaccine/Immunisation |

**Figure 1**

Predictive performance associated with calibrated models using isotonic calibration

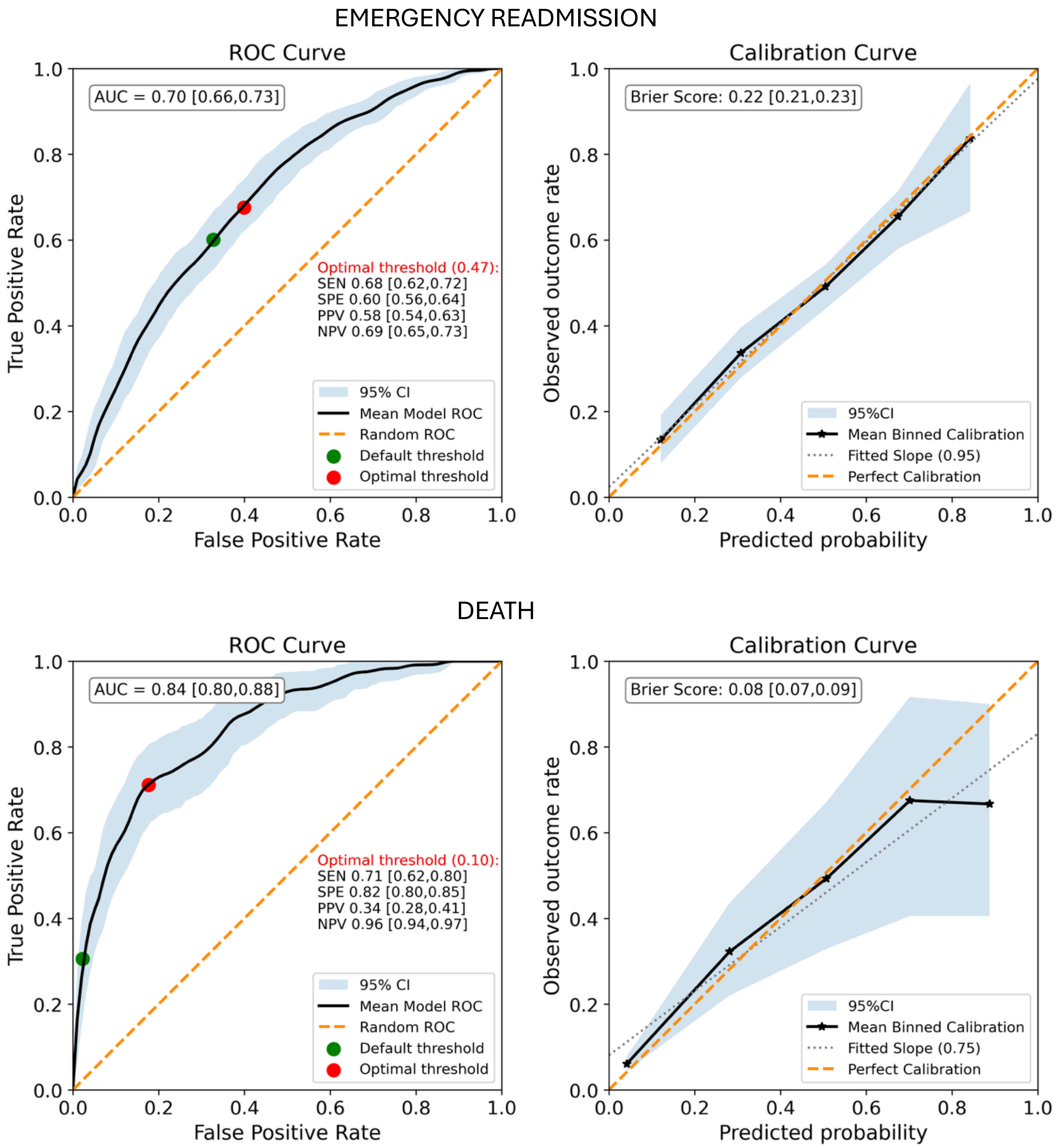

*Figure 1*: Receiver Operating Curves and calibration curves corresponding to the emergency readmission risk prediction model (top panel) and the mortality risk prediction model

### Completed TRIPOD+AI Checklist

*Paper: Explainable, Personalised Prediction of Emergency Readmission and Mortality Following Hospitalization in Patients with Heart Failure*

Notes: Page numbers refer to manuscript pages in the attached paper. NR = not reported. N/A = not applicable. “Partial” indicates the item is partly addressed but a TRIPOD+AI component is missing or only implicit.

| Section | Topic | Item | D/E | Checklist item | Reported on page | Completion note |
|---|---|---|---|---|---|---|
| TITLE | Title | 1 | D;E | Identify the study as developing or evaluating the performance of a multivariable prediction model, the target population, and the outcome to be predicted | 1 | Title identifies explainable personalised prediction of emergency readmission and mortality following HF hospitalisation. |
| ABSTRACT | Abstract | 2 | D;E | See TRIPOD+AI for Abstracts checklist | 1 | Structured abstract reports background, methods, results and conclusions. |
| INTRODUCTION | Background | 3a | D;E | Explain the healthcare context (including whether diagnostic or prognostic) and rationale for developing or evaluating the prediction model, including references to existing models | 2-3 | Prognostic context and rationale are described, including post-discharge adverse events, prior risk models and gaps. |
| INTRODUCTION | Background | 3b | D;E | Describe the target population and the intended purpose of the prediction model in the context of the care pathway, including its intended users | 3, 16 | Target population and purpose are reported; intended users are mainly implied as clinicians supporting post-discharge planning. |
| INTRODUCTION | Background | 3c | D;E | Describe any known health inequalities between sociodemographic groups | NR | Variables related to sociodemographic groups included sex, country of birth, language spoken at home, and Indigenous status. Health inequalities between Indigenous and non-Indigenous Australians are well documented. However, the present study was not designed or approved to undertake comparative analyses by Indigenous status. Such analyses require appropriate Indigenous governance and ethics approval, including review and oversight by Aboriginal and Torres Strait Islander representatives. Therefore, Indigenous status was considered as a candidate predictor, but subgroup comparisons of model performance or outcomes by Indigenous status were not undertaken. |
| INTRODUCTION | Objectives | 4 | D;E | Specify the study objectives, including whether the study describes the development or validation of a prediction model (or both) | 3 | Objectives specify development/evaluation of explainable patient-level risk stratification for 180-day emergency readmission and mortality. |
| METHODS - Data | Data | 5a | D;E | Describe the sources of data separately for the development and evaluation datasets, the rationale for using these data, and representativeness of the data | 3-4 | CardiacAI EMR linked to APDC and RBDM are described. Random train/test split uses same data source. Representativeness is not fully addressed. |
| METHODS - Data | Data | 5b | D;E | Specify dates of collected participant data, including | 3-4 | Study period reported as 1 Jan 2017 to 31 Mar 2021; |

| Section | Topic | Item | D/E | Checklist item | Reported on page | Completion note |
|---|---|---|---|---|---|---|
| | | | | participant accrual start/end and, if applicable, end of follow-up | | outcomes assessed within 180 days of discharge. |
| METHODS - Participants | Participants | 6a | D;E | Specify key elements of the study setting including the number and location of centres | 3-4 | Secondary care setting: seven hospitals in NSW, Australia; CardiacAI data from two Local Health Districts. |
| METHODS - Participants | Participants | 6b | D;E | Describe the eligibility criteria for study participants | 3-4 | Adults with HF diagnosis; exclusions include in-hospital death, transfers, hospice/EOL/palliative care, and LOS <24 hours. |
| METHODS - Participants | Participants | 6c | D;E | Give details of any treatments received, and how they were handled during model development or evaluation, if relevant | 5 | Discharge/current medication orders are handled as predictors, grouped using Cerner Multum DNUM and filtered by prevalence/relevance. |
| METHODS | Data preparation | 7 | D;E | Describe any data pre-processing and quality checking, including whether this was similar across relevant sociodemographic groups | 6 | Train/test partitioning, one-hot encoding, standardisation, binary mapping and feature filtering are described. Quality checking by sociodemographic group is not reported. |
| METHODS - Outcome | Outcome | 8a | D;E | Clearly define the outcome and time horizon, including how and when assessed, rationale, and consistency across sociodemographic groups | 3-4 | Outcomes are all-cause emergency readmission and all-cause death within 180 days, identified via APDC/RBDM. Rationale tied to STRONG-HF horizon. |
| METHODS - Outcome | Outcome | 8b | D;E | If outcome assessment requires subjective interpretation, describe qualifications and demographic characteristics of outcome assessors | N/A | Outcomes are derived from linked administrative/death registry data, not subjective assessment. |
| METHODS - Outcome | Outcome | 8c | D;E | Report any actions to blind assessment of the outcome to be predicted | 6 | Test data including outcomes to be predicted were not part of model development. |
| METHODS - Predictors | Predictors | 9a | D | Describe the choice of initial predictors and any pre-selection before model building | 4-6 | Initial predictors cover demographics, social/family history, admission characteristics, prior utilisation, pathology, vital signs, medications, problems, procedures and text-derived symptoms; filtering/pre-selection criteria are reported. |
| METHODS - Predictors | Predictors | 9b | D;E | Clearly define all predictors, including how and when measured | 4-6 | Predictor groups and timing/measurement summaries are reported. Full variable list appears to rely partly on supplementary material. |
| METHODS - Predictors | Predictors | 9c | D;E | If predictor measurement requires subjective interpretation, describe qualifications and demographic characteristics of predictor assessors | 4-6 | Most predictors are extracted from EMR/administrative data; text concepts use rule-based extraction. These keyword search is not assessed. The number of mentions is taken as a proxy for importance. |
| METHODS | Sample size | 10 | D;E | Explain how study size was arrived at and justify sufficiency, including any sample size calculation | 7,18 | Cohort derivation flowchart is reported and small sample size is discussed. |

| Section | Topic | Item | D/E | Checklist item | Reported on page | Completion note |
|---|---|---|---|---|---|---|
| METHODS | Missing data | 11 | D;E | Describe how missing data were handled. Provide reasons for omitting any data | 4-6, 17 | Missingness thresholds, exclusions and handling of missing values are described. |
| METHODS - Analytical methods | Analytical methods | 12a | D | Describe how the data were used for model development/evaluation, including partitioning | 6 | Data were randomly partitioned 80/20 at patient level and stratified by outcome. |
| METHODS - Analytical methods | Analytical methods | 12b | D | Describe how predictors were handled in the analyses | 5-6 | Categorical variables one-hot encoded; continuous variables standardised; binary variables mapped to 0/1; missingness left unimputed for tree-based modelling. |
| METHODS - Analytical methods | Analytical methods | 12c | D | Specify model type, rationale, model-building steps, hyperparameter tuning, and internal validation | 6 | XGBoost gradient-boosted decision trees; Optuna Bayesian optimisation; patient-stratified cross-validation; bootstrap-resampled ensemble; isotonic calibration. |
| METHODS - Analytical methods | Analytical methods | 12d | D;E | Describe if/how heterogeneity in parameter estimates/performance was handled across clusters | 6-7 | Bootstrap resampling/prediction intervals quantify uncertainty. |
| METHODS - Analytical methods | Analytical methods | 12e | D;E | Specify all measures and plots used to evaluate model performance | 6, 8, 11 | ROC curves, AUC, Brier score, calibration curve and slope are reported; threshold-based PPV/sensitivity/specificity are also shown. |
| METHODS - Analytical methods | Analytical methods | 12f | E | Describe any model updating arising from model evaluation | 6-7 | Isotonic calibration was applied and reportedly had small calibration improvement without significant performance improvement. |
| METHODS - Analytical methods | Analytical methods | 12g | E | For model evaluation, describe how model predictions were calculated | 6-7 | Predictions are ensemble averages across bootstrap models; SHAP explanations are averaged across bootstrapped training models. |
| METHODS | Class imbalance | 13 | D;E | If class imbalance methods were used, state why/how and any recalibration | N/A | No class imbalance method was used. |
| METHODS | Fairness | 14 | D;E | Describe approaches used to address model fairness and rationale | N/A | Fairness was not the focus of this study. |
| METHODS | Model output | 15 | D | Specify output of prediction model and details/rationale for classification thresholds | 6, 8-9 | Model outputs are predicted probabilities/risk scores. Classification threshold uses maximum geometric mean of sensitivity and specificity; alternative lower threshold is illustrated for readmission. |
| METHODS | Training versus evaluation | 16 | D;E | Identify differences between development and evaluation data in setting, eligibility, outcome and predictors | 6 | Development/test sets are random patient-level partitions from the same cohort. |
| METHODS | Ethical approval | 17 | D;E | Name ethics committee/IRB and describe participant consent or waiver | 17 | Ethics committees and approval numbers are reported. CardiacAI operates under an opt-out model whereby posters are displayed in participating hospitals and information sheets are made available to patients on request. |

| Section | Topic | Item | D/E | Checklist item | Reported on page | Completion note |
|---|---|---|---|---|---|---|
| OPEN SCIENCE | Funding | 18a | D;E | Give source of funding and role of funders | 17 | MRFF Cardiovascular Health Mission Grant funding is reported. Funders' role is not explicitly described, although authors state views are their own. |
| OPEN SCIENCE | Conflicts of interest | 18b | D;E | Declare conflicts of interest and financial disclosures for all authors | 17 | Authors have no conflict of interest. |
| OPEN SCIENCE | Protocol | 18c | D;E | Indicate where the study protocol can be accessed or state that no protocol was prepared | N/A | |
| OPEN SCIENCE | Registration | 18d | D;E | Provide registration information or state the study was not registered | N/A | |
| OPEN SCIENCE | Data sharing | 18e | D;E | Provide details of availability of study data | 18 | Information on data access can be found at cardiacai.org |
| OPEN SCIENCE | Code sharing | 18f | D;E | Provide details of availability of analytical code | 18 | Information on code access will be provided following acceptance in peer-reviewed journal. |
| PATIENT & PUBLIC INVOLVEMENT | Patient & Public Involvement | 19 | D;E | Provide details of patient/public involvement or state no involvement | 18 | Information on stakeholder involvement can be found at cardiacai.org |
| RESULTS | Participants | 20a | D;E | Describe participant flow, numbers with/without outcome, follow-up time; diagram helpful | 7-8 | Cohort derivation flowchart/table and 30-day/180-day outcome counts are reported. |
| RESULTS | Participants | 20b | D;E | Report characteristics overall and where applicable for each data source/setting, including key dates, predictors, treatments, sample size, outcome events, follow-up, missing data and demographic differences | 8-10 | Baseline characteristics are summarised by outcome, with selected predictors and medication classes. Detailed missing data/demographic subgroup differences are limited. |
| RESULTS | Participants | 20c | E | For model evaluation, compare with development data distribution of important predictors, demographics and outcome | N/A | Not required given the nature of the train/test partition. |
| RESULTS | Model development | 21 | D;E | Specify number of participants and outcome events in each analysis | 7-9 | Overall cohort and outcome event counts are reported; test-set threshold counts are reported. Counts for tuning/resampling are not fully specified. |
| RESULTS | Model specification | 22 | D | Provide details of full prediction model to allow predictions in new individuals and third-party evaluation/implementation | 5-7 | Model can be replicated using standard libraries: XGBoost/Optuna/SHAP |
| RESULTS | Model performance | 23a | D;E | Report performance estimates with confidence intervals, including for key subgroups; plots aid presentation | 8-11 | AUC and Brier score with intervals, calibration slopes/curves and ROC curves are reported. Subgroup performance is not reported. |
| RESULTS | Model performance | 23b | D;E | If examined, report heterogeneity in model performance across clusters | N/A | Not examined/reported. The sample size is too small for inspection of model performance across clusters. |
| RESULTS | Model updating | 24 | E | Report results from model updating, including updated model and subsequent performance | N/A | |
| DISCUSSION | Interpretation | 25 | D;E | Give overall interpretation of main results, including fairness in | 15-17 | Main results are interpreted against previous studies. Fairness is not discussed. |

| Section | Topic | Item | D/E | Checklist item | Reported on page | Completion note |
|---|---|---|---|---|---|---|
| | | | | context of objectives and previous studies | | |
| DISCUSSION | Limitations | 26 | D;E | Discuss limitations and effects on bias, uncertainty and generalisability | 17 | Limitations include modest sample size, need for external validation, text-extraction limitations, medication capture and competing risks. |
| DISCUSSION | Usability in current care | 27a | D | Describe how poor quality/unavailable input data should be assessed and handled when implementing the model | N/A | Limitations are described in the discussion section. This model is not yet intended for implementation. Future work includes larger cohorts, external validation, improved NLP/LLMs and time-to-event/competing-risk analyses. |
| DISCUSSION | Usability in current care | 27b | D | Specify whether users must interact in input data handling/use of model and required expertise | N/A | Limitations are described in the discussion section. This model is not yet intended for implementation. Future work includes larger cohorts, external validation, improved NLP/LLMs and time-to-event/competing-risk analyses. |
| DISCUSSION | Usability in current care | 27c | D;E | Discuss next steps for future research with a view to applicability and generalisability | 16-17 | Future work includes larger cohorts, external validation, improved NLP/LLMs and time-to-event/competing-risk analyses. |
